# Expected endpoints from future chikungunya vaccine trial sites informed by serological data and modeling

**DOI:** 10.1101/2022.05.23.22275466

**Authors:** Quan Minh Tran, James Soda, Amir Siraj, Sean Moore, Hannah Clapham, T. Alex Perkins

**Author notes:** 100 Galvin Life Science Center, Notre Dame, IN 46556.

## Abstract

In recent decades, there has been an increased interest in developing a vaccine for chikungunya. However, due to its unpredictable transmission, planning for a chikungunya vaccine trial is challenging. To inform decision making on the selection of sites for a vaccine efficacy trial, we developed a new framework for projecting the expected number of endpoint events at a given site. In this framework, we first accounted for population immunity using serological data collated from a systematic review and used it to estimate parameters related to the timing and size of past outbreaks, as predicted by an SIR transmission model. Then, we used that model to project the infection attack rate of a hypothetical future outbreak, in the event that one were to occur at the time of a future trial. This informed projections of how many endpoint events could be expected if a trial were to take place at that site. Our results suggest that some sites may have sufficient transmission potential and susceptibility to support future vaccine trials, in the event that an outbreak were to occur at those sites. In general, we conclude that sites that have experienced outbreaks within the past 10 years may be poorer targets for chikungunya vaccine efficacy trials in the near future. Our framework also generates projections of the numbers of endpoint events by age, which could inform study participant recruitment efforts.

## Introduction

Chikugunya virus (CHIKV) is an arthropod-borne virus (arbovirus) that is transmitted by mosquitoes. It is estimated that 1.3 billion people are currently living in areas at risk [1]. The first documented global emergence of the virus started from Tanzania in 1952 and led to subsequent outbreaks in sub-Saharan Africa, India, and Southeast Asia [2,3]. Though reports of chikungunya cases diminished after its emergence in the 1950s, it has recently expanded to the Americas and Europe [4–6]. The rapid spread and long-term effects of CHIKV [7–11] highlight the need to find ways to improve on prevention of these outcomes. Like most viral infections, there is no specific treatment for chikungunya, and patient care mostly relies on supportive treatment. This emphasizes the need for prevention. Though vector control has been widely used for arboviruses, it is difficult to employ effectively and often has only modest impacts [12–14]. Thus, vaccination still remains the most promising prevention strategy when a safe and efficacious vaccine is available.

Although no vaccines for chikungunya are currently licensed, there are several vaccine candidates in development. The vaccines in phase I or phase II have shown promising results with highly immunogenic reactions and minimal adverse effects [15]. Current vaccine candidates in phase III relied on neutralizing antibody titers as the endpoints [16,17]. Since this surrogate of protection is still debatable, additional vaccine trials that focus on clinical endpoint events are still needed to truly assess the efficacy of these vaccine candidates. During public health emergencies, vaccine candidates can be licensed before phase-III trials completed under the World Health Organization (WHO) prequalification program. This has been the case for an Ebola vaccine, and most recently for COVID-19 vaccines [18,19]. However, chikungunya has not proceeded under this program. Thus, completing phase-III clinical trials is necessary for the vaccine candidates to be licensed and become widely available for the public.

Vaccine development for CHIKV faces a serious obstacle when it comes to demonstrating efficacy [15]. This owes to the fact that it is notoriously difficult to predict when and where chikungunya outbreaks will occur due to the sporadic epidemiology of this disease. This means that vaccine trials most likely need to be conducted during one or more outbreaks [20]. The considerable logistical difficulties associated with conducting a trial during an outbreak place even more importance on being prepared to carry out trials in locations where outbreaks may be likely. Continuing outbreaks of chikungunya globally [21] offer encouragement about the prospect of efficacy trials being possible, although current vaccine candidates have so far not been able to leverage these outbreaks as opportunities for phase-III efficacy trials required for licensure. Completion of a phase-III vaccine efficacy trial depends not just on a chikungunya outbreak occurring somewhere, but in a particular population that can be enrolled and followed during a trial. Advancements in the ability to appropriately select sites with a high probability of a future outbreak are needed to speed up the evaluation of these vaccine candidates.

Mathematical modeling is playing an increasingly important role in vaccine trial planning in general [22], including in the problem of site selection [23–25]. Site selection is a major challenge for chikungunya vaccine efficacy trial planning due to its sporadic epidemiology. This problem is traditionally solved with the help of epidemiological data to estimate recent incidence of the proposed efficacy endpoint, such as reported cases of symptomatic disease. For chikungunya, incidence is dynamic and not fixed over time. Moreover, it is likely that past chikungunya outbreaks may have gone unreported or misdiagnosed [26]. Analysis of age-stratified serological data offers one way to reconstruct recent patterns of CHIKV transmission relevant to vaccine efficacy trial planning [27,28], but such analyses have only been performed in a limited number of settings and have not been leveraged to directly inform vaccine efficacy trial planning.

In this study, we use age-stratified serological data and mathematical modeling to project vaccine trial endpoint incidence in the event of a future outbreak. Our approach consists of three steps: 1) infer epidemic parameters from age-stratified serological data; 2) determine the population immunity; and 3) project outbreak size and endpoint events if an outbreak were to occur at the time of a vaccine trial. We first demonstrate our framework in a simulation study, with the purpose of understanding the accuracy of our inferences when the true values of the underlying parameters are known. We then apply this approach to 35 sites for which age-stratified serological data were available, although we also propose this as a method that could be applied to age-stratified serological data collected as part of pre-trial epidemiological studies.

## Materials and methods

### Inform vaccine efficacy trial endpoints from serological data

For a given location, we 1) estimate the basic reproduction number *R*_0_, the timing of past outbreaks *y*, and average initial susceptible proportion before the first outbreak happened *S*_0_(0) from serological data, and 2) determine population immunity at the time of data collection. Assuming a hypothetical outbreak happens during a vaccine efficacy trial in 2022, we then 3) project the *IAR* of the outbreak and the number of endpoints expected among a given number of subjects. This framework is summarized visually in Fig. 1. All of the code for this analysis was written in the R language [29] and is available, along with all data required to reproduce this study, in a GitHub repository [30].

**Figure 1.**
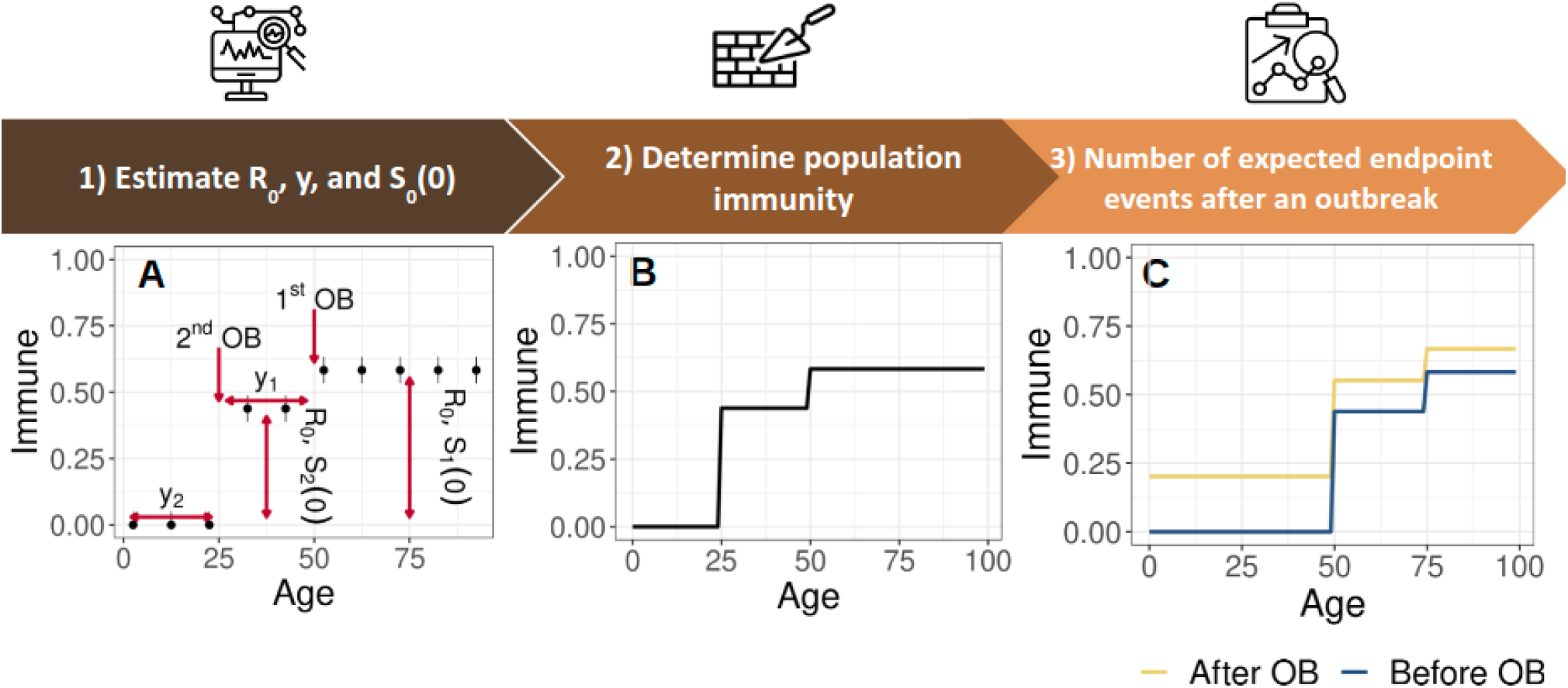
Framework to project the number of endpoint events in a future vaccine trial. Our framework consists of three stages: 1) using serological data available before the vaccine trial, we can estimate the basic reproduction number *R*_0_, the average initial susceptible proportion before the first outbreak *S*_0_(0) (used to inform the susceptible proportion before the first outbreak *S*_1_(0) (Supplementary Text)), and the timing of past outbreaks *y* (panel A). 2) From the estimated parameters in A, we can derive the population immunity during the serological study at the site (panel B). 3) The population immunity at the time of a future vaccine trial can then be projected during the vaccine trial (panel C). In the event that an outbreak happens during that future trial, we project the infection attack rate (*IAR*) of the outbreak using the estimated *R*_0_ and the projected population immunity. The number of expected endpoint events can be derived subsequently. Note: *S*_1_(0) and *S*_2_(0) are the susceptible proportion before the first and second outbreaks respectively; *y*_1_ and *y*_2_ are the timing of the first and second outbreaks; OB: outbreak

#### 1. Estimate R_0_, y, and S_0_(0) from age-stratified serological data

We assumed that the number positive *p*_*i*_ in each age group *i* follows a binomial distribution, such that the log likelihood informed by serological data is

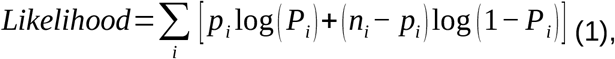

where *n*_*i*_ is the number of samples and *P*_*i*_ is the model’s prediction of the proportion positive. We can derive *P*_*i*_ from the age-stratified susceptibility in the population *S*_*a,Y*_ in the year *Y* and age *a* at which the data were collected, which equals

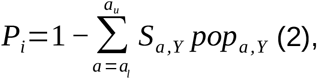

where *a*_*l*_ and *a*_*u*_ are the lower bound and upper bound of age group *i* and *pop*_*a,Y*_ is the proportion of people in age group *a* at year *Y*. This age-specific proportion was derived from annual population by age data from United Nations World Population Prospects [31], assuming the site has the same age demographic as in country level.

Consistent with the sporadic nature of chikungunya outbreaks, we assume that the age-stratified susceptibility *S*_*a,Y*_ has a step-like shape, where each “step” of the seroprevalence curve corresponds to an outbreak in the past [32]. The particular shape can be modeled in our framework with an SIR model in which the years of past outbreaks are specified, along with the basic reproduction number *R*_0_ and the population susceptibility prior to the first outbreak within the lifetime of the oldest study participants *S*_0_(0). The time between outbreaks corresponds to the width of each step, while the susceptible-infected-recovered (SIR) model informs the proportion immune following each outbreak, which corresponds to the height of each step (Fig. 1A). Details on how to derive *S*_*a,Y*_ for given values of *R*_0_, the initial susceptible proportion, and the timing of past outbreaks *y* are described in the Supplementary Text.

For each seroprevalence data set, we fitted five models, which differed in terms of the number of historical outbreaks (1 to 5 outbreaks). We estimated the parameters *R*_0_, *S*_0_(0), and *y* in a Bayesian framework, using the *BayesianTools* package [33]. We used the DREAMzs algorithm, with one million iterations and a burn-in period of 500,000 iterations. The assumed prior distributions for the parameters were

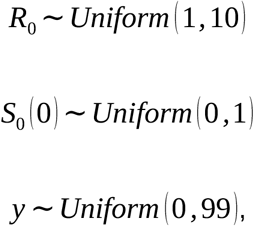

with the latter two being non-informative and the range for the *R*_0_ prior being defined broadly within the range of values generally observed for chikungunya [34]. Convergence was assessed by calculating the potential scale reduction statistic 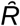 using the *coda* package in R [35]. We applied a post-processing reversible-jump Markov chain Monte Carlo algorithm from the *rjmcmc* package [36] to derive ensemble weights of the five models for each data set. These weights were then used to generate ensemble predictions.

#### 2. Determine population seropositivity at the time of the study

From the estimates of *R*_0_, *y*, and *S*_0_(0), we can reconstruct the step-like shape of population immunity in every age group at the time of the study. This reconstruction was done based on the relationship between the basic reproduction number (*R*_0_), the proportion susceptible in a population before an outbreak *S*(0), and the *IAR* over the course of an outbreak from a SIR model as stated in step 1 and in the Supplementary Text.

#### 3. Number of expected endpoint events after a hypothetical outbreak in 2022

For the next step, we projected the age-stratified positive proportion in the population to year 2022 to derive the positive proportion in the population before a hypothetical outbreak in 2022. The age-stratified positive proportion was estimated individually from each of the five models with different numbers of outbreaks.

Given the age-stratified positive proportion in the population in 2022 derived from model *m*, we can derive the infection attack rate for each model *IAR*_*m*_ if there was a hypothetical outbreak in 2022 based on equation (1) in Supplement Text. The ensemble prediction of *IAR* can be derived from the *IAR*_*m*_ for each model *m* and from the model weights estimated from the RJMCMC algorithm in Step 1, according to

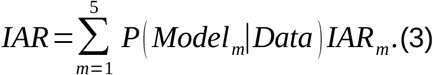

Consistent with the endpoint for chikungunya vaccine trials proposed by WHO [37], we chose symptomatic cases as the endpoint for the hypothetical vaccine trials in 2022 that we focused on in our analysis. The number of symptomatic cases among trial participants is generated by multiplying the predicted *IAR* by the proportion of infections that result in symptoms, which is sampled from a beta distribution with shape and scale parameters of 21.80 and 3.18, respectively. We derived this distribution from empirical estimates of the asymptomatic proportion of CHIKV infections ranging from 3% to 28% [38–40], such that 95% of samples would fall within this range.

### Simulation study

#### Model behavior across a range of parameter values

To assess the behavior of our model, we calculated the *IAR* during a hypothetical outbreak in 2022 across a range of parameter values. The range of *R*_0_ values was chosen to be from 1 to 3, which is the typical *R*_0_ values estimated from previous outbreaks [41]. We assume the study was conducted in 2000 and that past outbreaks may have happened between 1900 to 2000 in increments of ten years. We also explore how *IAR* will change if there is only one or multiple past outbreaks. We expect that *S*_0_(0) < 1 would have the same effect as an additional outbreak before the first one, which is already covered by exploring how multiple outbreaks affect *IAR* as just mentioned. Therefore, for simplicity, *S*_0_(0) was set to 1 so there was no immunity in the population before the first outbreak.

#### Performance of our inference method on simulated data

To assess the performance of our model at estimating parameters, we applied our inference method to simulated data from hypothetical serological studies with known parameters. These hypothetical studies involved 2,000 simulated study participants, stratified into 20 age groups of five years each. The outbreak times *y* were set to be 5, 45, and 85 years before the study time. For these simulations, each outbreak had *R*_0_ = 1.5 and *S*_0_(0) = 1. These simulated datasets were used to estimate back the parameters *R*_0_, *y*, and *S*_0_(0), which were then used to project the *IAR* during a hypothetical outbreak in 2022. To assess the performance of our approach when data is more limited, we also conducted a sensitivity analysis by changing the sample size (only ten participants for each of 20 age groups) or the age bins (only five age groups of 20 participants each), but keeping everything else the same. The age distribution of Kenya [31] was used in all of these simulations.

### Applying to published serosurveys

After validating our method on simulated data, we applied it to published data, which were obtained from a systematic review of CHIKV serological data [42]. Criteria for inclusion of studies in this review were that a study should report data from immunoglobulin G (IgG) enzyme-linked immunoassay (ELISA) test, plaque reduction neutralization tests (PRNT), or hemaglutination inhibition (HI) on at least three age groups collected as part of a cross-sectional study. We used estimates of population by age appropriate to the country and year of the study as estimated by the United Nations World Population Prospects [31].

## Results

### Simulation study

#### Model behavior across a range of parameter values

To understand the relationships between the model’s parameters and the predicted *IAR*, we derived the possible values of *IAR* for a hypothetical outbreak in 2022, given a range of possible values of *R*_0_ and the timing of past outbreaks. In the case of a site with only one outbreak, we observed that a longer time since the last outbreak and larger *R*_0_ would be associated with a higher *IAR* in 2022 (Fig. 2). When the time since the last outbreak was less than 10 years, the *IAR* was less than 5%, regardless of the *R*_0_ value.

**Figure 2.**
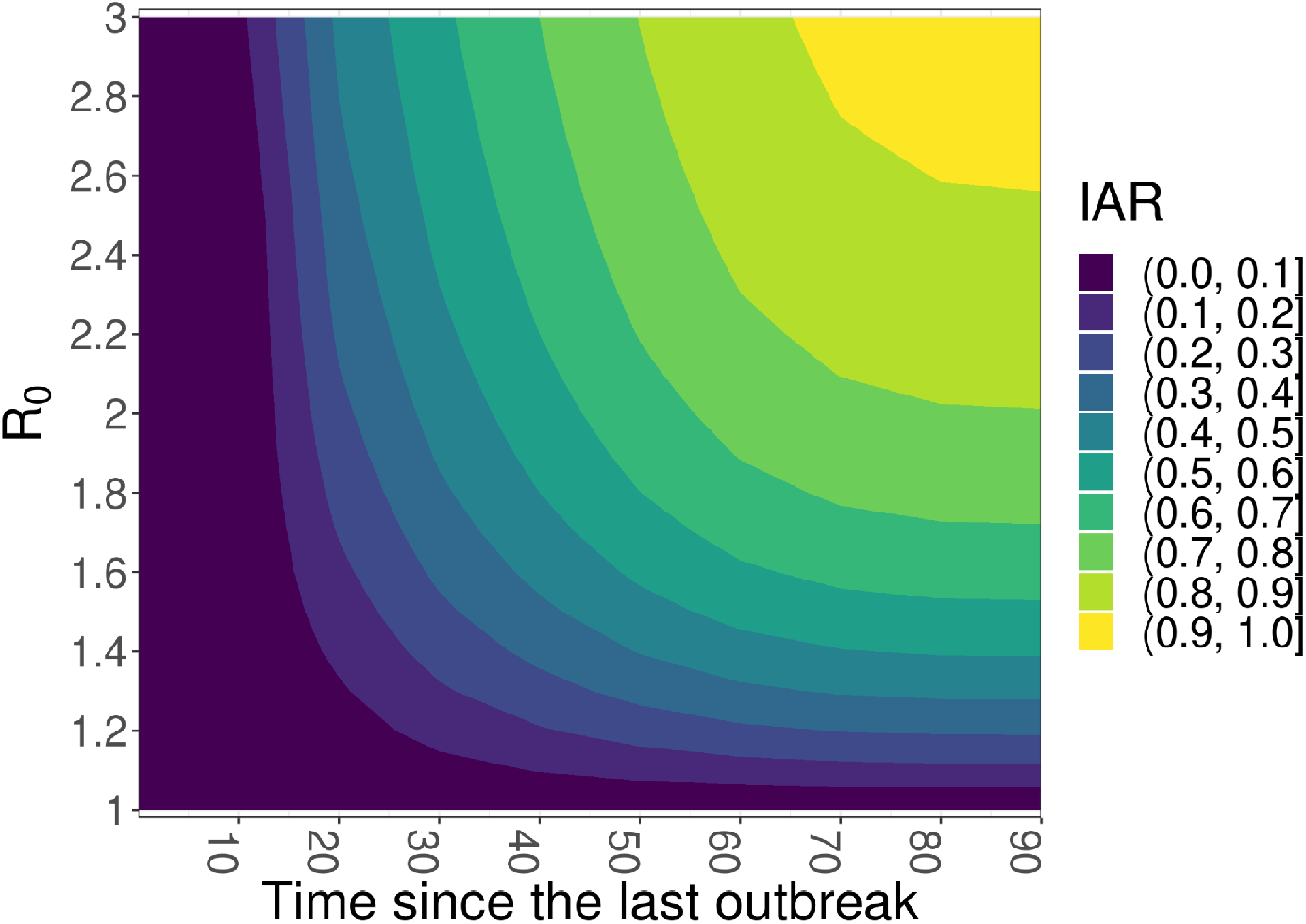
The relationship between *R*_*0*_, time from the last outbreak results in *IAR* in the future outbreak in the model with only one outbreak. The magnitude of the future *IAR* generated is colored as shown in the legend. We observed that in the model with one outbreak, higher *R*_0_ and the further the time from the last outbreak will result in higher *IAR*.

When there have been multiple outbreaks at a site, the relationship between *R*_0_ and *IAR* is more complicated (Fig. S1, Fig. S2). We observed that as the time since the last outbreak and the value of *R*_0_ increase, the average values of *IAR* tend to increase (Fig. S1). This relationship is similar to what we observed when we simulated data on a single outbreak. However, as the time between the past outbreaks increases, the average values of *IAR* tend to decrease (Fig. S2). This occurs because as the latest outbreaks are further away, a higher susceptible proportion builds up between outbreaks. The subsequent outbreak then depletes this susceptible proportion and leaves a smaller susceptible pool for the future hypothetical outbreak. This will eventually lead to a decrease in the final *IAR* of the future hypothetical outbreak. For example, if the most recent outbreak was in 2000, it would have been larger if the outbreak before that had been in 1970 than if it had been in 1990. Thus, an outbreak in 2022 would be smaller because the most recent outbreak in 2000 had been larger.

#### Performance of our inference method on simulated data

To evaluate the performance of our model for estimating parameters and predicting *IAR*, we applied it to simulated data on age-stratified seroprevalence with predefined parameter values of *R*_0_ = 1.5, *S*_0_(0) = 1, and outbreak times of 5, 45, and 85 years before the serological study. Our reconstruction of age-stratified seroprevalence, as expected, exhibited three step-like increases corresponding to three past outbreaks (Fig. S4). The model probabilities for the five models placed the most weight on the model with three outbreaks (94.8%, Fig. S3). Overall, the uncertainty intervals of our estimates covered the true values well (Fig. S3). Furthermore, the ensemble posterior prediction from the five models produced a good prediction to the age-stratified serological data (Fig. S4).

We did assess our model performance on simulated data with more outbreaks (not presented here) but found out that with *R*_0_ = 1.5, more than three outbreaks within 100 years depleted enough susceptibles that the final outbreak sizes became minimal and unable to be inferred.

To assess how well the inference framework performs on less ideal data, we carried out a sensitivity analysis in which we reduced the sample size or increased the size of the age bins (Figs. S5-S8). Overall, this led to more uncertainty about the number of past outbreaks. That notwithstanding, the estimates of *R*_0_, *S*_0_(0), and the timing of past outbreaks covered the simulated values and fit the data reasonably well for models close to the correct number of past outbreaks.

Since estimation of *R*_0_ and the projection of the proportion susceptible at the time of a future vaccine trial have the greatest influence on *IAR*, correctly identifying the number of past outbreaks may not be strictly necessary if population susceptibility can still be ascertained. To explore this possibility, we compared *IAR* values projected using true parameter values versus parameter values inferred from different simulated data sets. When we used data sets with smaller sample sizes and larger age bins, we found that the projected *IAR* values were comparable to the true *IAR* values in terms of their central tendency, but more uncertain in comparison to the more detailed simulated data set (Fig. S9).

### Applying the model to published serosurveys

We applied our framework to age-stratified seroprevalence data identified through a literature review [42]. In total, we used data from 18 studies consisting of 35 site-level datasets, given that some studies reported data from multiple sites. A detailed description of each study is available in the Supplementary Text and the colated data can be found in [dataset][30]. The study sites were labeled alphabetically from A to R for convenience.

#### Parameter estimates

All of the model runs converged according to the potential scale reduction statistic 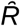. Estimates of *R*_0_ for all studies were within the range of 1 to 3 (Fig. 3), which is typical for chikungunya [41]. Most sites had model weights for the one-outbreak model of >90%, showing strong support for a one-outbreak model. However, several sites showed more support for a model with two outbreaks (Fig. 3; sites A, H1, J, I2, I4, I5, I7, I8, L2), and one site in the Philippines (M) appears to have had five outbreaks (model weight of five-outbreaks model is 99.5%). Estimates of *S*_0_(0) for most sites were highly uncertain, which is to be expected given that most information in the data about susceptibility pertained to the period in which study participants were alive, rather than before then.

**Figure 3.**
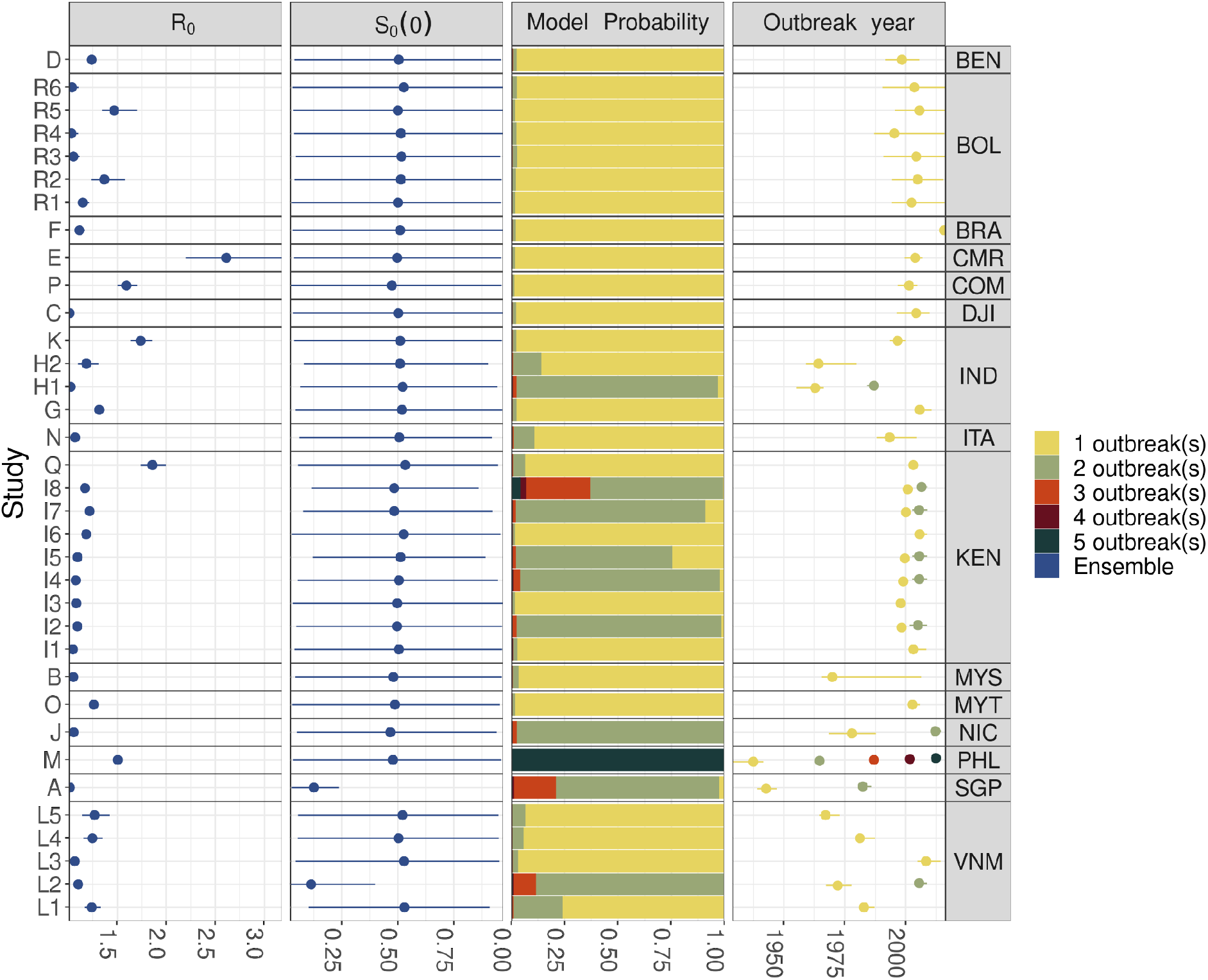
Parameter estimates for every study site. Points and ranges in each row belong to one study site, representing the median and 95% Crl estimates of the parameters respectively. The first and second column show the ensemble distribution (blue) of *S*_0_(0) and *R*_0_ from the five models. The third column shows the model weight for each of the five models. The fourth column is the timing of outbreak(s) estimated from the model that has the largest weight. The right labels are the ISO-3 code of the country where the site was conducted. Table S1 and Fig. S11 provide the code of the study site.

#### Expected number of endpoint events

Utilizing the estimated parameters, we computed the population immunity of the population in 2022 before and after a hypothetical outbreak (Fig. S11), and subsequently projected the expected number of endpoint events for every site. The numbers of endpoint events were projected to be highly variable across sites. Only two sites (L5 and E) were projected to have the 2.5 quantile of expected endpoint events higher than ten events per thousand study participants (Fig. 4). These two sites either had higher *R*_0_ (site E) or higher susceptibility in the population before the hypothetical outbreak (site L5) (Fig. 4). When susceptibility was high, increasing *R*_0_ increased the expected number of events. However, when susceptibility was low, a much higher *R*_0_ value was required to result in an equivalent number of events, as observed for site E (Fig. 4). A positive interaction between *R*_0_ and susceptibility was also observed when we inspected the relationship between the time since the last estimated outbreak and the number of events (Fig. 5). With high enough *R*_0_, site E is expected to still obtain a higher number of events despite more recently experiencing an outbreak.

**Figure 4.**
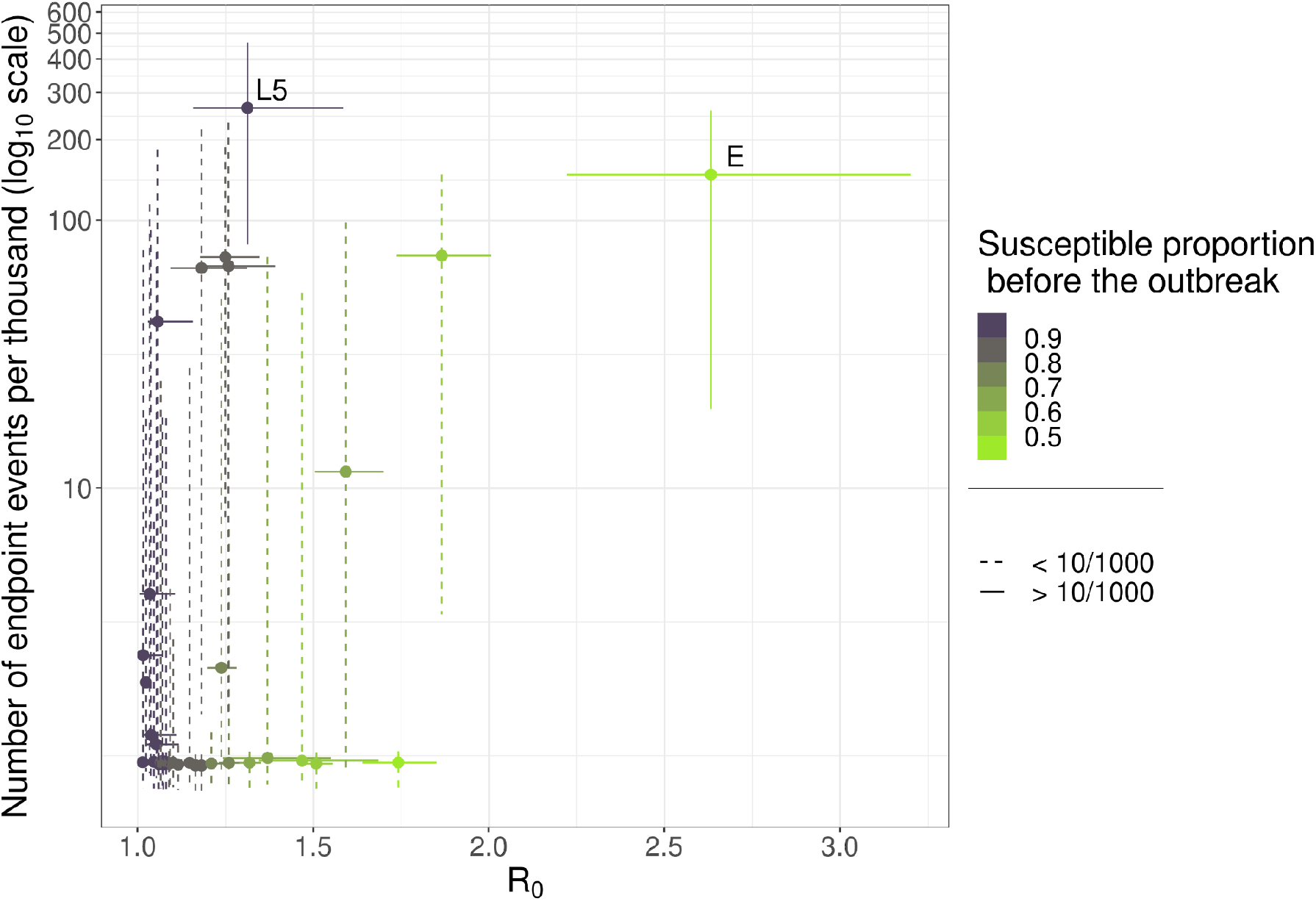
Relationship between *R*_0_, estimated number of endpoint events, and susceptible proportion before a hypothetical outbreak in 2022 for every site. Each point and range are the median and 95% PPI for each site. The dashed lines represent studies that have at least part of the 95% PPI below the value of 10 per 1,000 people. We annotated study sites that were confidently projected to produce more endpoints than that in the event of an outbreak.

**Figure 5.**
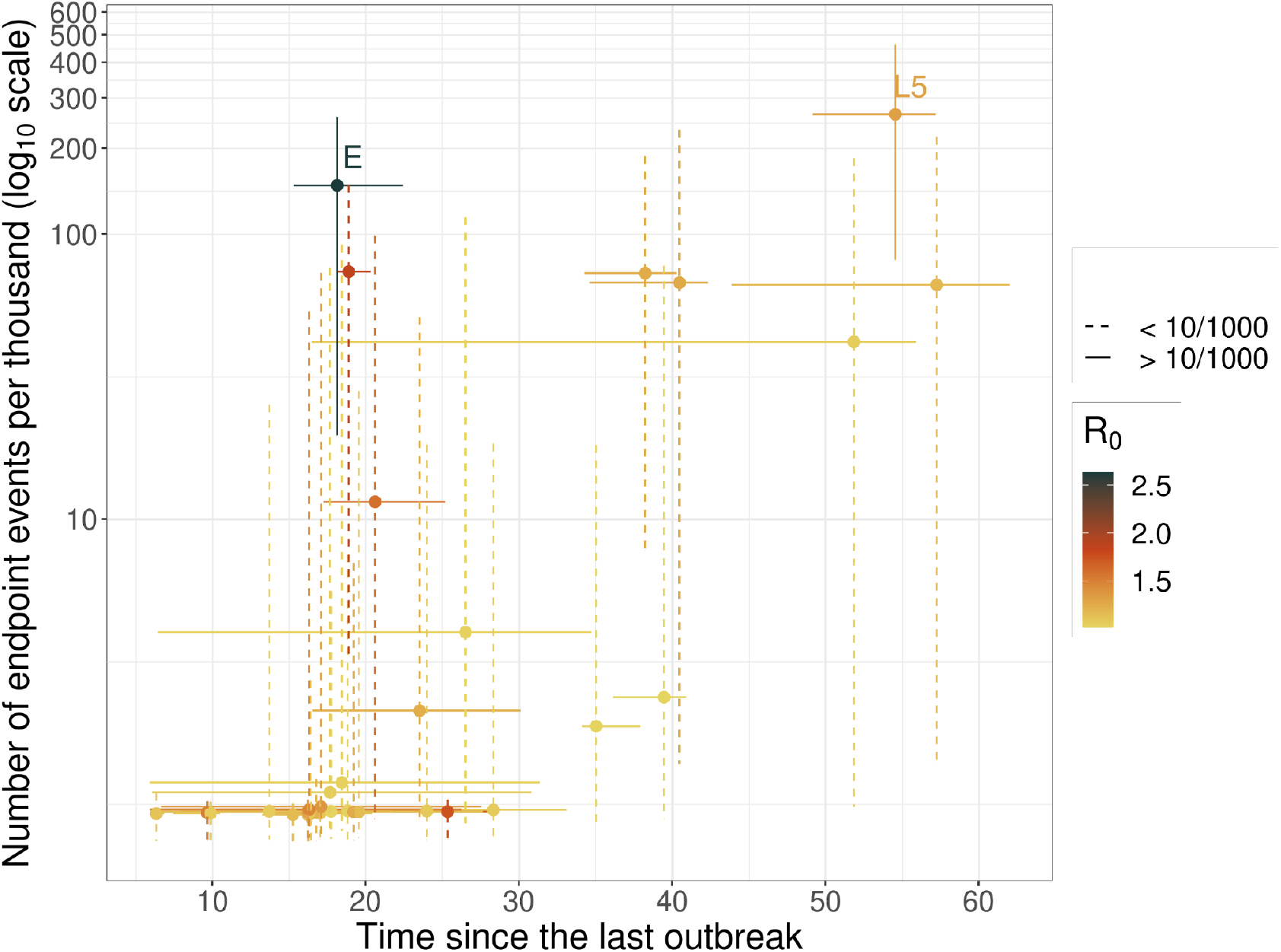
Relationship between time since the last outbreak, estimated number of endpoint events, and *R*_0_ for every site. Each point and range are the median and 95% PPI for each site. The number of expected endpoints was plotted in *log*_10_ scale. The dash linerange represents studies that have the 95% PPI cross or below the value of 10 per 1000 people. We annotated study sites that can confidently produce more than 10 endpoints per thousand people.

Because historical outbreaks can affect the population immunity of older and younger generations differently, we also stratified our results into two age groups—children (<18 years old) and adults (≥18 years old)—and compared results between them. Adults generally have experienced more outbreaks in the past than children; hence, susceptible proportions before hypothetical outbreaks in 2022 are expected to be higher in children. This results in a higher expected number of events in children than adults (Fig. 6).

**Figure 6.**
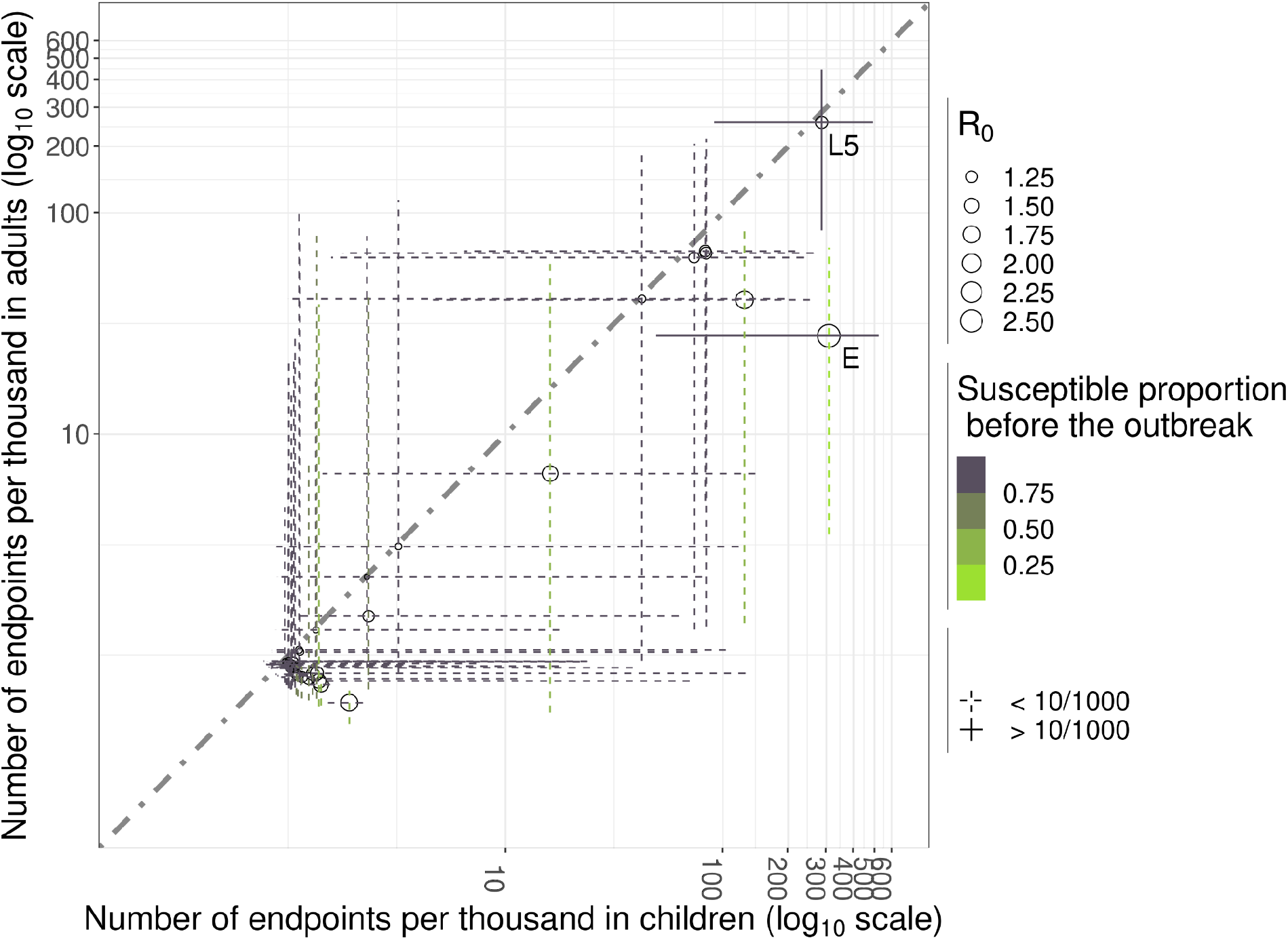
Comparing the number of expected events from a hypothetical outbreak in 2022 in different target groups: adults and children. Each point and range are the median and 95% PPI for each site. The number of expected endpoints was plotted in *log*_10_ scale. The dash linerange are sites that have the 95% PPI of the number of endpoints crossing or below the value of 10 per 1000 people. The size of each point refers to the magnitude of estimated *R*_0_ from each site. We annotated study site that can produce more than 10 endpoints per thousand people

### Validation

We performed an additional analysis on site M at which the serological surveys were conducted in multiple years (1973, 2012, and 2013). We used parameters *R*_0_ and *S*_0_(0) estimated from the serological survey in 1973 to project seropositive proportions from every age group following outbreaks in 2012 and 2013. We compared those projections with the positive proportions of every age group estimated from fitting to data from all years. The same estimated timings of past outbreaks after 1973 were used for both the predictions. We estimated *R*_0_ from the data in 1973 to be 1.22 (95% CrI: 1.13 - 1.33), which was lower than an estimate of 1.51 (95% CrI: 1.46 - 1.55) from the data from all years. Although the model run on 1973 data fit well to the serological data on the year (Fig. 7A), the lower seropositive proportion predicted in 1973, and most importantly the lower estimate of *R*_0_, led to a lower projection of the seropositive proportion in older age groups in 2012 and 2013 (Fig. 7B, 7C). The model runs that included data from all years resulted in a good fit for the serological data in 2012 and 2013 (Fig 7E and 7F). However, the higher *R*_0_ estimate from this model run resulted in a poorer fit to the 1973 serological data (Fig. 7D). Consistent with these results, the *IAR* projections right after the five previous outbreaks in the site using only the 1973 dataset were also lower (Fig. 8). This reflects that there may be variability in the *R*_*0*_ over time that our model does not capture.

**Figure 7.**
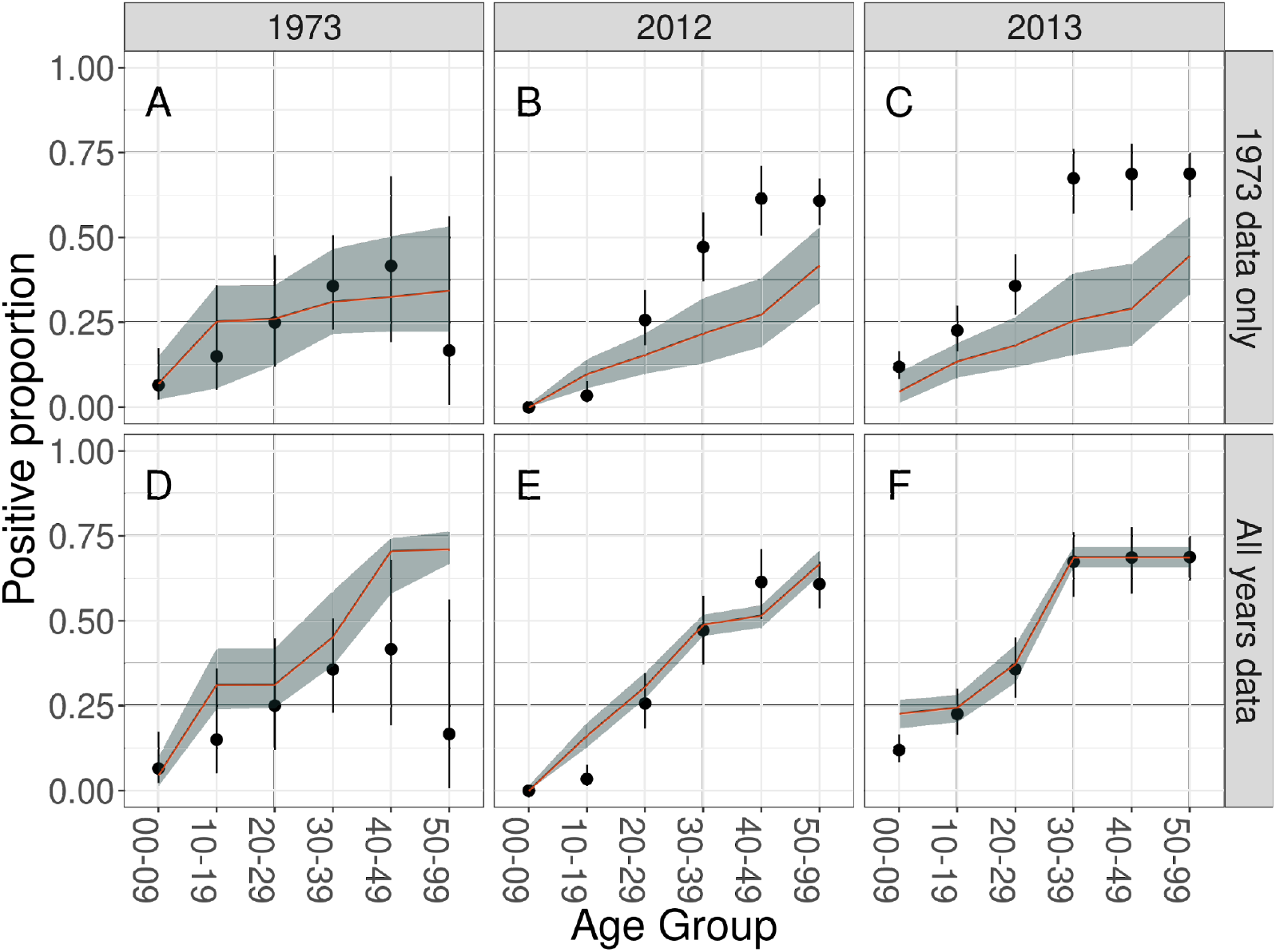
Posterior predictions of seropositivity for serosurveys conducted in 1973, 2012, and 2013 att site M in the Philippines. Panel A is the posterior prediction of 1973 serosurvey using *R*_0_, *S*_0_(0), and the timing of past outbreaks estimated from the model with two outbreaks but applying for the 1973 data only. Panel B and C are the datafit generated using *R*_0_, *S*_0_(0) from the 1973 data but timing of past outbreaks estimated from the data from all years. Panels D, E, and F are the posterior prediction for every serosurvey using parameter estimates from the model with five outbreaks for the data from all years. The points with their ranges show the true values and 95% PPI of the positive proportion for every age group in the simulation data. The red line and the gray area are the median and 95% PPI of ensemble positive proportion for every age group estimated from the five models.

**Figure 8.**
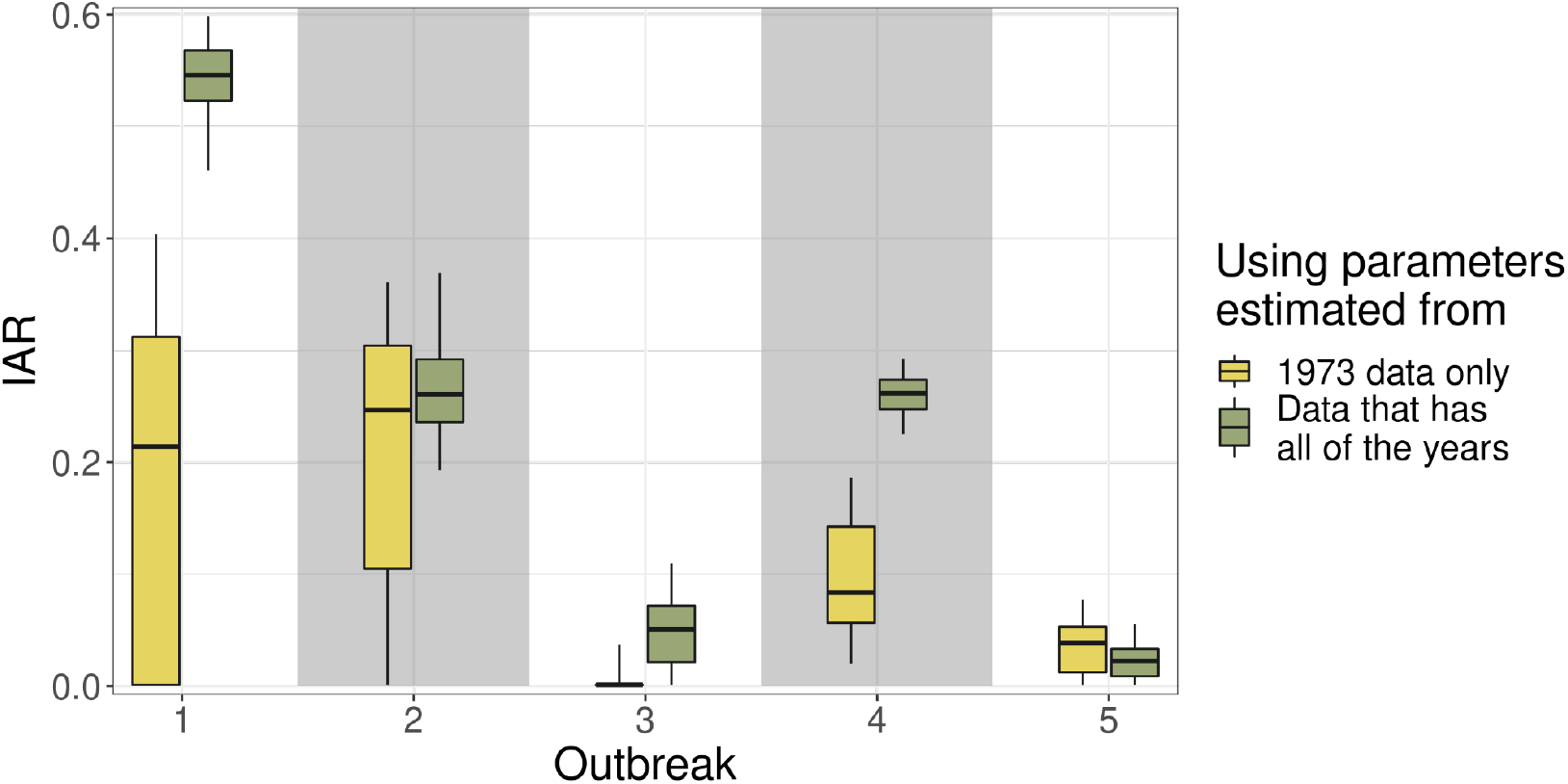
Infection attack rate of the previous five outbreaks for the Philippines informed by data from different combinations of years. The boxplots are the *IAR* projected using different data sets: the first two outbreaks that use estimates from 1973 data only (outbreak 1 and 2 in yellow), the three latest outbreaks that use estimates from 1973 data but with the timing of past outbreaks estimated from the data that use all of the years (outbreak 3 to 5 in yellow), and the five outbreaks that use estimates from the data from all years (green).

We also compared our estimates of the timing of past outbreaks to the timing of reported outbreaks (Table S1) not used to inform our estimates (Fig. 9). Overall, most of our estimates match the timing of reported outbreaks. However, the model performed less well for studies A (Singapore) and K (Kerala, India). This is due to the fact that these studies only sampled patients older than 18 years for study A and older than 14 years for study K. Missing information from younger age groups may result in the model picking up the signal from unreported earlier outbreaks instead. Interestingly, data sets from study R (Bolivia) and D (Benin) also apply to the same age bins, yet our model still captured the timing of those reported outbreaks within its credible intervals, though with a high uncertainty. This discrepancy is because these sites may have had only one recent outbreak happen at any time between when the oldest of the missing reported person was born and the time the study was conducted. Our model cannot pinpoint exactly the timing of these outbreaks from these missing ranges, thus explaining the high uncertainty in the estimates. The studies from I1 to I8 were conducted in different places in Kenya; hence, there may be some variation in the estimates of the timing of past outbreaks.

**Figure 9.**
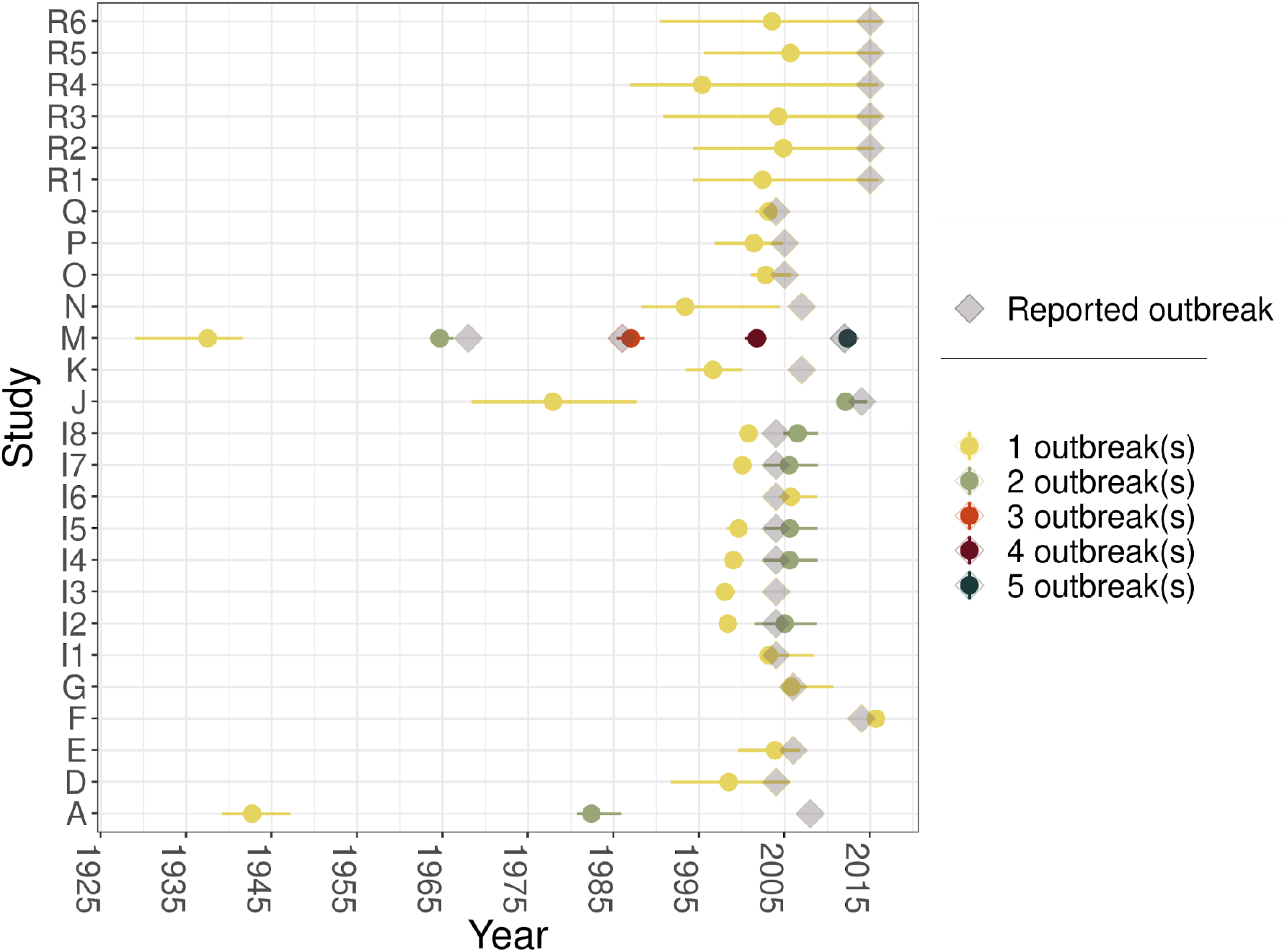
The timing of past outbreak estimates from our model as compared to the timing of reported outbreaks. For every study (coded as shown in Table S1 or Figure S11), each point and range is the estimated median and 95% CrI of the timing of past outbreaks from our model. The number of previous outbreaks are color-coded as in the legend. The diamond shapes are the reported timing of outbreaks at each of those sites.

## Discussion

In this study, we constructed a framework to make projections of expected endpoints for future vaccine trials based on serological data for a candidate trial site. The framework employs the historical transmission information imprinted in serological data to infer the susceptibility of the population and a time-averaged *R*_0_ from previous outbreaks. These quantities can then be used to make projections of *IAR* for future outbreaks. We first applied this framework to simulated data to assess its performance and then applied it to published serosurveys to make inferences about hypothetical future trials. We propose that this framework is suitable for planning a vaccine trial conducted during an outbreak and is independent of any particular trial design, as all trials need endpoint events to be successful.

Our results suggest that, in most cases, outbreaks may be minimal if the time since the last outbreak is 10 years or less, even at relatively high *R*_0_ values. This is because it takes time for the susceptible population to build up through births to a sufficient degree for an outbreak to happen. This fits with previous reports of CHIKV outbreaks in some places, where it may take 10 or more years to re-emerge in a given population [43–45]. Similar expectations have been described for Zika, another arbovirus with similar epidemiology [46].

When applying this framework to published serological data, our estimates of the timing of past outbreaks generally agree with the timing of reported outbreaks at the same site. Although estimates at some sites were not as good due to missing samples in younger age groups, this can be noted and improved by collecting data for all ages in preparation for future vaccine trials. Most of the sites we analyzed here appear to have experienced only a single outbreak within the lifetimes of study participants, which is consistent with the idea that the occurrence of chikungunya outbreaks is sporadic. We inferred that the site in Cebu City, Philippines appeared to have experienced multiple outbreaks, a conclusion reached by a previous analysis using the same data set but using a slightly different method [28].

Based on a WHO consultation for future phase-III chikungunya vaccine trials [37], future clinical trials to assess vaccine efficacy need to be conducted across multiple sites and multiple countries due to the unpredictable nature of CHIKV outbreaks [37]. These sites also need to be chosen prospectively in a way that they accrue enough endpoint events that the trial has enough power to detect a desired efficacy. Our framework can be used to support this objective by offering a basis for determining which sites are expected to generate a larger number of endpoint events, in the event that an outbreak occurs. Based on the aforementioned WHO consultation, 62 endpoint events are needed to have 90% power to reject the null hypothesis that the vaccine efficacy is less than 30% when the true efficacy is 70% [37]. Our analysis identified two sites that alone could potentially accrue the required 62 endpoint events with fewer than 6,200 study participants if an outbreak were to happen. One additional suggestion from our framework is to potentially target enrollment on individuals who were born after past outbreaks, given that they would be more likely to be susceptible.

Our approach addresses only one criterion for site selection: the expected incidence of endpoint events. Conducting a vaccine trial during an outbreak poses other challenges that the site selection process needs to take into account. A suitable site should also have trained personnel, regulatory oversight, and testing facilities. There are also questions on how many sites are needed and allocation of effort across sites. To answer this, a balance between the expected incidence of endpoint events and limited resources is necessary. Many sites across multiple countries also require considerable logistical effort to communicate among sites. These challenges have been noted in a previous Ebola vaccine trial [47]. Ultimately, selecting suitable sites for chikungunya vaccine efficacy trials is a multifaceted problem and the modeling framework we proposed here can act as a screening tool so that the sites it recommends can be further scrutinized according to other criteria, as well.

Our study has some limitations. For an arbovirus like CHIKV, heterogeneity plays an important role in transmission, which could affect the relationship between *R*_0_ and the *IAR* [48,49]. However, estimation of R0 based on observed IAR values is not the end goal of our method. Rather, the goal here is to having a model that is capable of using past values of IAR and an estimate of R0 to predict future IAR values. Thus, the omission of heterogeneity may lead to an inaccurate estimate of R0 but does not necessarily compromise our main goal to project future IAR [48,49]. Because we assume that *R*_0_ for every outbreak is the same, our framework will not perform as well in situations in which there is variation in transmission between outbreaks [50]. This could partly explain why the *R*_0_ estimates in the validation for the Philippines site are different and lead to different projections of *IAR*. Analyzing seroprevalence data from too far back could be a potential problem since we may miss recent outbreaks. This could be remedied by conducting pilot seroprevalence studies, which is usually done before the vaccine trial anyway. Another limitation of our analysis is that the symptomatic proportion is sampled from a distribution derived from a widely cited asymptomatic proportion ranging 3 - 28% [38–40]. However, there is growing evidence that the asymptomatic proportion of CHIKV may be lineage dependent [51]. This could easily be modified in our framework when the circulating lineage in the selected site for the trial is known, or at least suspected. It is also worth noting that our framework does not attempt to suggest when and where future outbreaks will occur, but instead to assess how feasible a vaccine trial would be if an outbreak occurs. Predicting the occurrence and timing of future chikungunya outbreaks remains a challenge and is beyond the scope of this study.

In conclusion, we have established a comprehensive framework that can estimate the number of expected endpoint events to inform future chikungunya vaccine efficacy trials using age-stratified serological data. Our analysis shows that this framework is robust when applied to simulated data but can also provide insights when applied to published data. In addition to chikungunya, this framework could also be applied to other emerging viruses with similar epidemiological dynamics.

## Data Availability

All data produced are available online at github

https://github.com/tranquanc123/CHIKV_endpoint_est.git

## Abbreviations

CHIKV: Chikugunya virus
WHO: World Health Organization
IAR: Infection attack rate
SIR: susceptible-infected-recovered
IgG: immunoglobulin G
ELISA: enzyme-linked immunoassay
PRNT: plaque reduction neutralization tests
HI: hemaglutination inhibition

## Fundings

This research was supported by a Young Faculty Award from the Defense Advanced Research Projects Agency (D16AP00114) and Start up grant from Saw Swee Hock School of Public Health.

## Conflict of interest

TAP receives consulting fees and research support from Emergent Biosolutions for work on chikungunya vaccine trial planning. The study described in this manuscript was completed prior to TAP’s relationship with Emergent.

## Notes

### Author Declarations

The source data is openly available to anyone beforehand. We used serological data from this systematic review: https://doi.org/10.1371/journal.pntd.0006533

