## Supplement Figures for "Expected endpoints from future chikungunya vaccine trial sites informed by serological data and modeling"

### Supplemental figures

#### Infection attack rate when there are multiple outbreaks

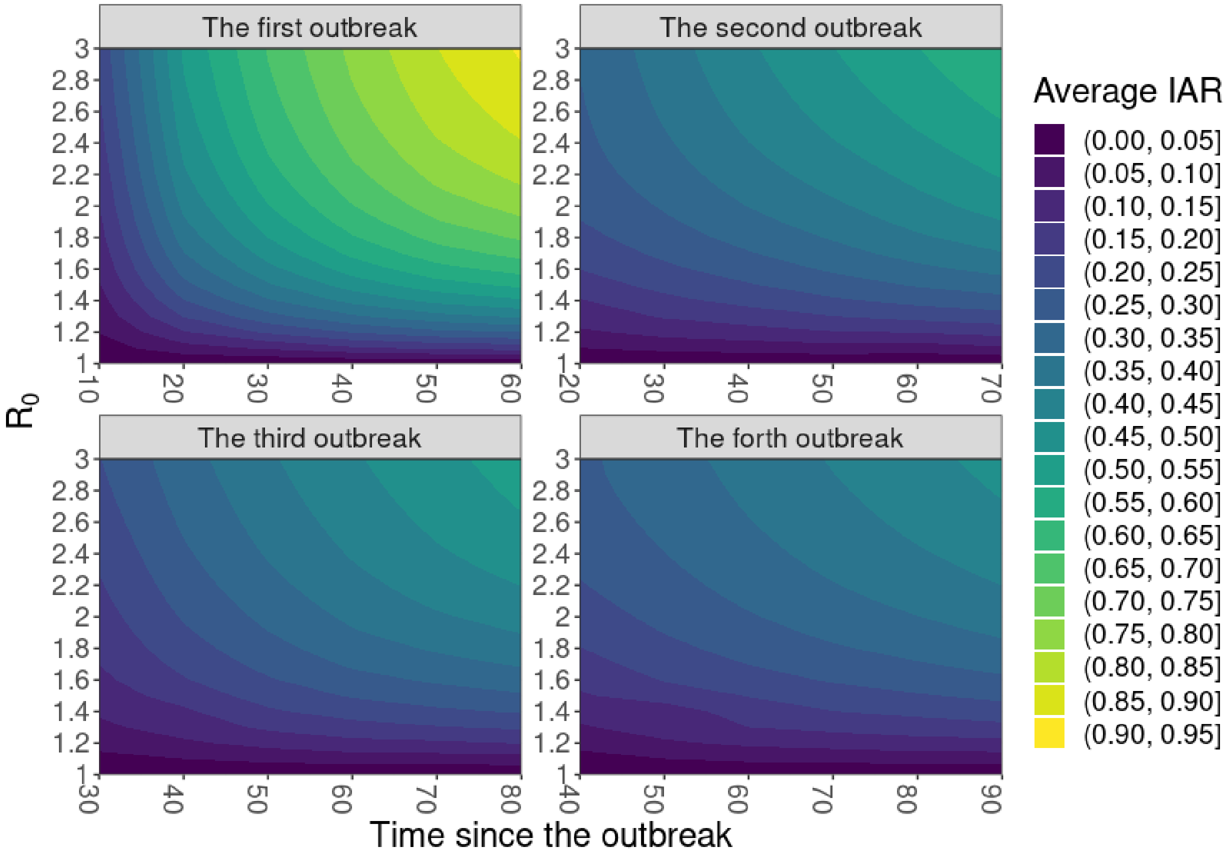


**Figure S1. The relationship between $R_0$, timing since the last outbreak and the average infection attack rate of the future outbreak in the simulated data with four outbreaks.** The magnitude of the average *IAR* of the future outbreak is colored as shown in the legend. As observed from the graph, the average values of *IAR* get bigger as the time since the outbreaks are larger and the value of $R_0$ is larger. This relationship is what we also observed in the simulated data with one outbreak as stated in the main text.

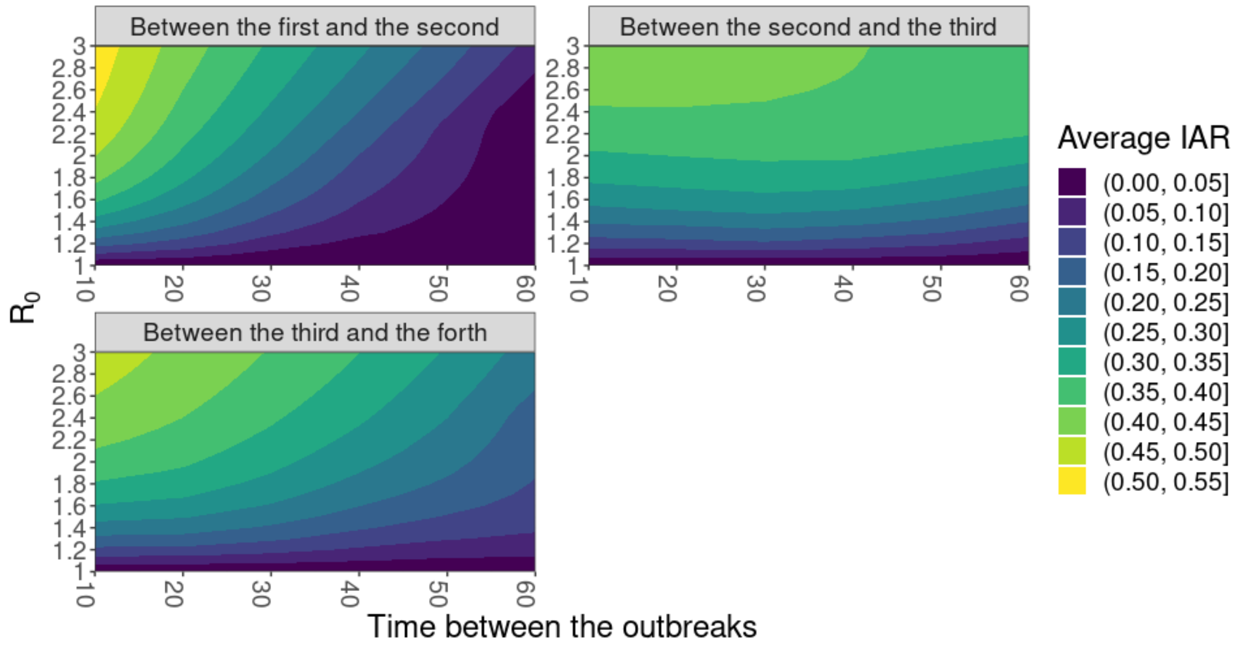


**Figure S2. The relationship between $R_0$, time between the outbreaks and the average infection attack rate of the future outbreak in the simulated data with four outbreaks.** The magnitude of the average *IAR* of the future outbreak is colored as shown in the legend. As observed from the graph, the average values of *IAR* get bigger as the time since the outbreaks are smaller and the value of $R_0$ is larger.

### Inference from simulated data

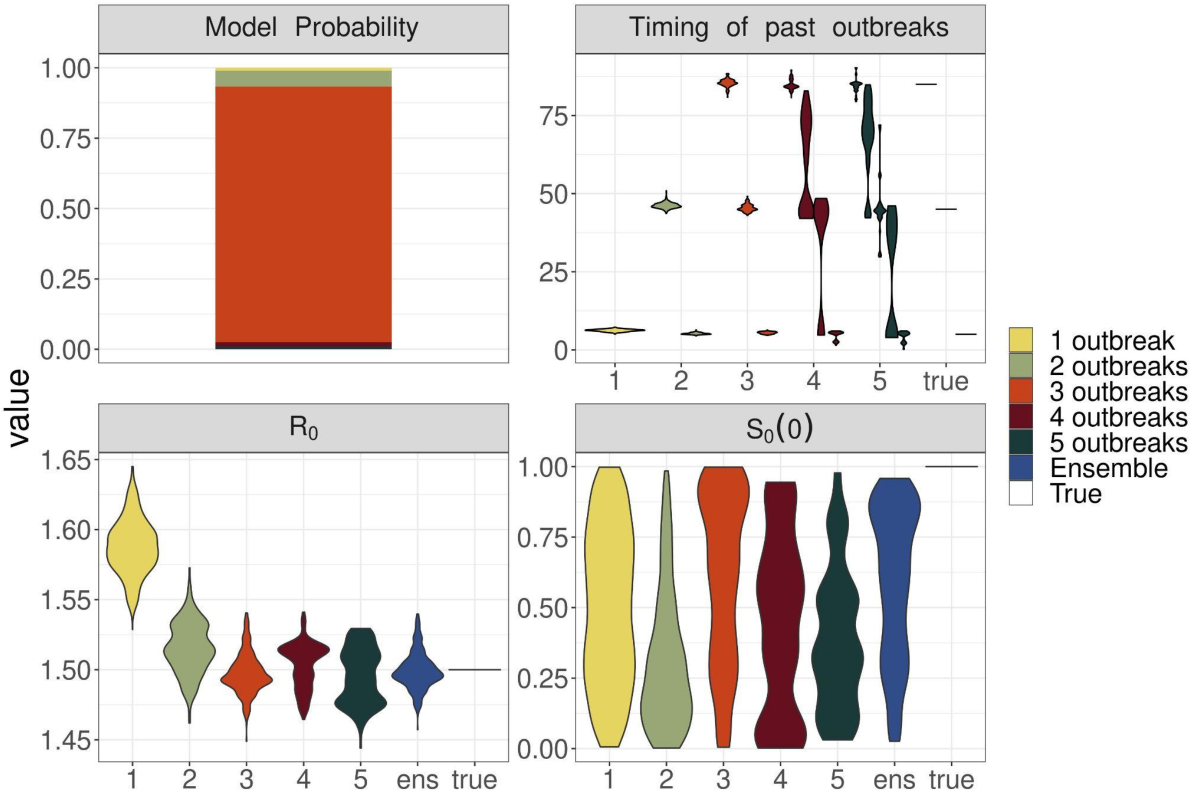


**Figure S3. Parameters estimated from the simulation study**. The upper left panel is a stacked bar plot showing the weight of models with different numbers of outbreaks estimated from the RJMCMC algorithm. Other panels show the distribution of outbreak times, $S_0(0)$, and $R_0$ estimated from each of the five models with different numbers of outbreak. The ensemble distribution (blue) of $S_0(0)$ and $R_0$ from the five models covers the true value of the parameters (horizontal lines in each panel).

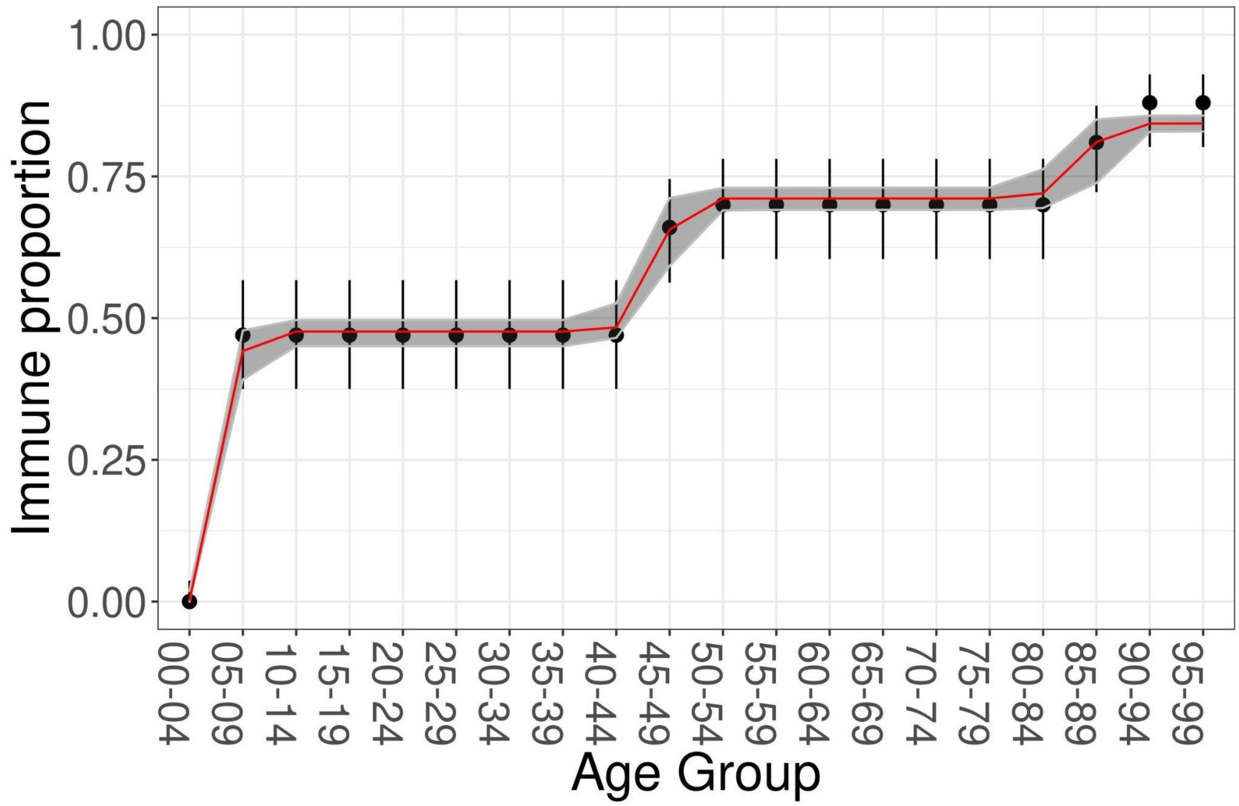


**Figure S4. Simulated age-stratified serological data and the posterior prediction of seropositivity from the five models**. The points with their ranges show the true values and 95% confident interval (CI) of the positive proportion for every age group in the simulation data. The red line and the gray area are the median and 95% posterior prediction interval (PPI) of ensemble positive proportion for every age group estimated from the five models.

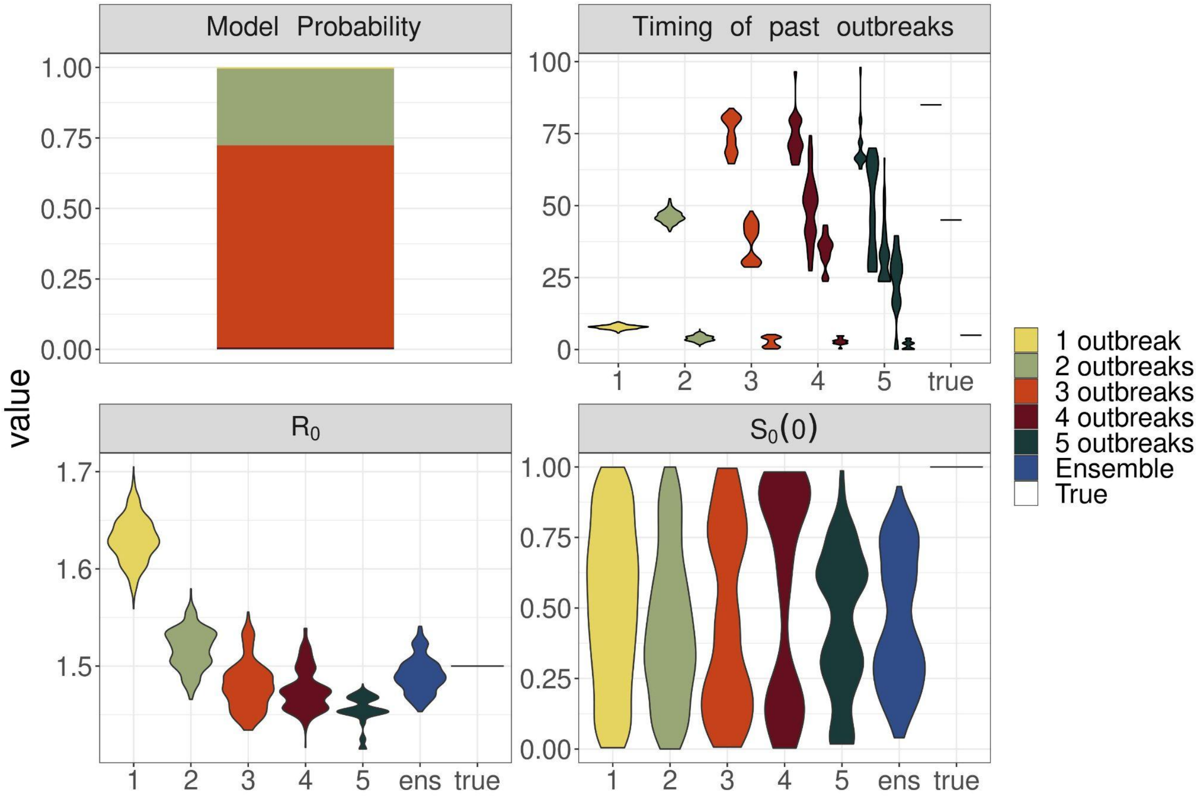


**Figure S5. Parameters estimated from the simulation study with larger age bins (age groups of 20)**. The upper left panel is a stacked bar plot showing the weight of models with different numbers of outbreak estimated from the RJMCMC algorithm. Other panels show the distribution of outbreak times, $S_0(0)$, and $R_0$ estimated from each of the 5 models with different numbers of outbreak. The ensemble distribution (blue) of $R_0$ from the 5 models covers the true value of the parameters (horizontal lines in each panel).

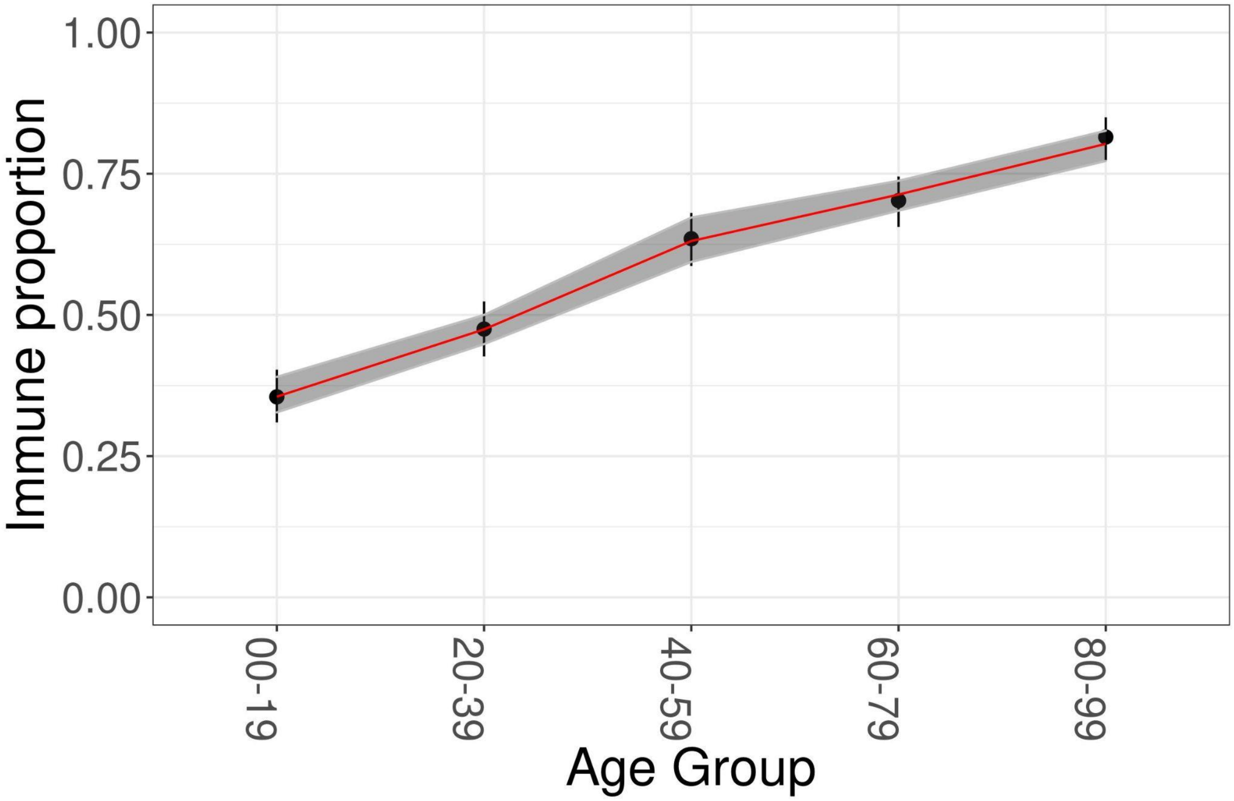


**Figure S6. Simulated age-stratified serological data with larger age bins (age groups of 20) and the posterior prediction of seropositivity from the five models**. The points with their ranges show the true values and 95% CI of the positive proportion for every age group in the simulation data. The red line and the gray area are the median and 95% PPI of ensemble positive proportion for every age group estimated from the five models.

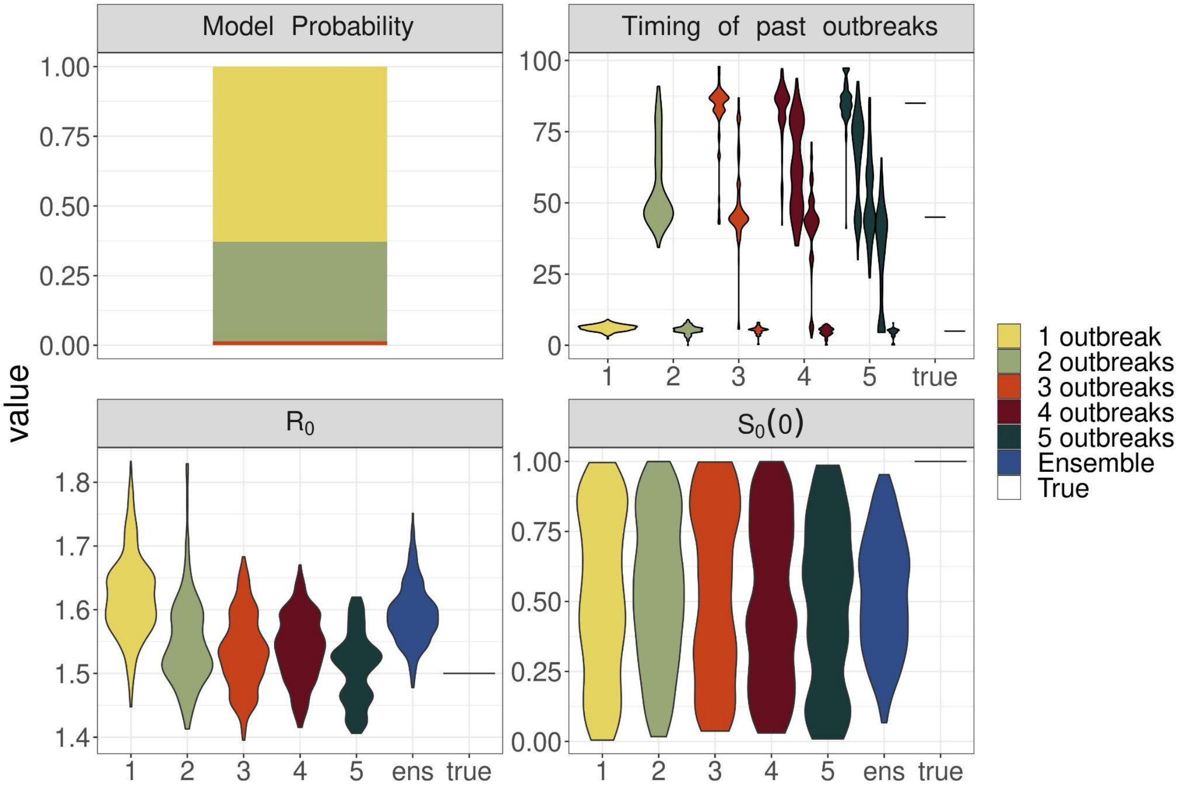


**Figure S7. Parameters estimated from the simulation study with less samples per age group (10 samples per age group)**. The upper left panel is a stacked bar plot showing the weight of models with different numbers of outbreak estimated from the RJMCMC algorithm. Other panels show the distribution of outbreak times, $S_0(0)$, and $R_0$ estimated from each of the 5 models with different numbers of outbreak. The ensemble distribution (blue) of $R_0$ from the 5 models covers the true value of the parameters (horizontal lines in each panel).

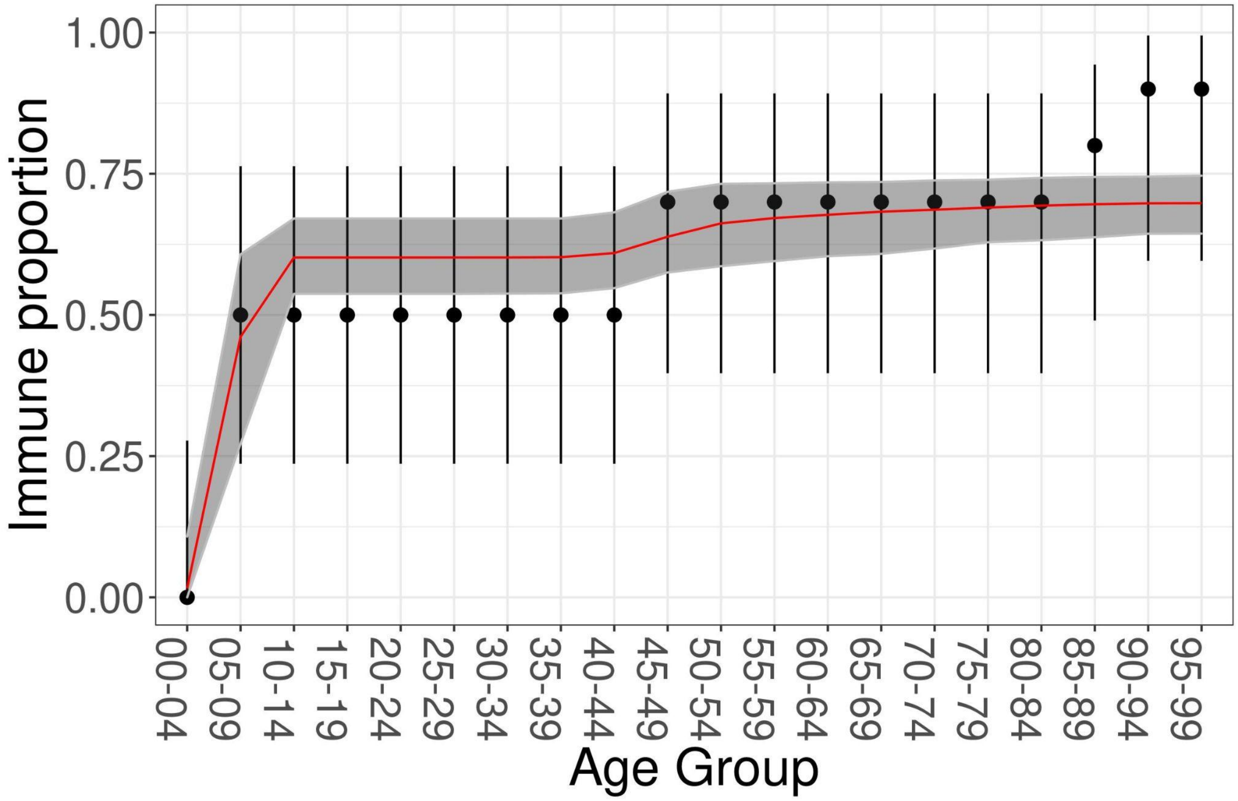


**Figure S8. Simulated age-stratified serological data with less samples per age group (10 samples per age group) and the posterior prediction of seropositivity from the five models**. The points with their ranges show the true values and 95% CI of the positive proportion for every age group in the simulation data. The red line and the gray area are the median and 95% PPI of ensemble positive proportion for every age group estimated from the five models.

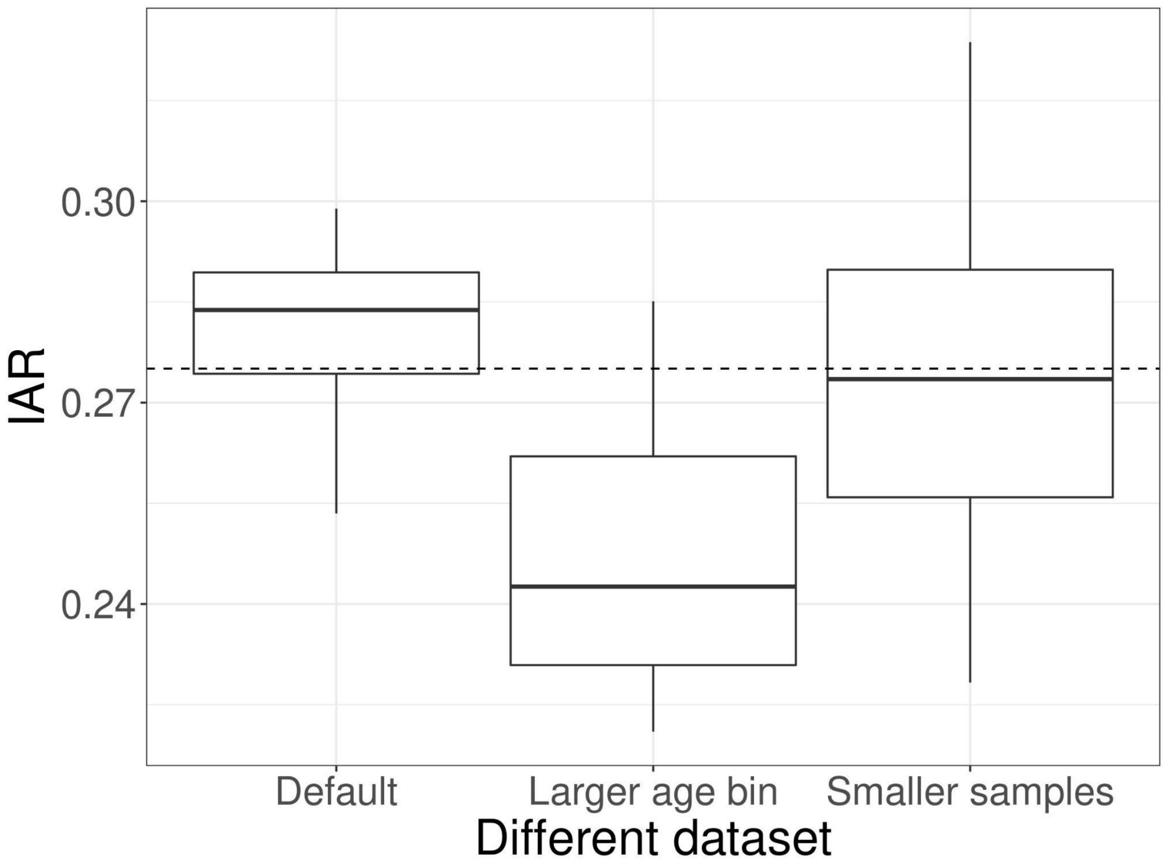


**Figure S9. Infection attack rate of a future outbreak projected based on model fits from different simulated data sets**. The first is from the default settings for simulated data, the data set with larger age bins (age group of 20), and the data set with 10 times smaller sample size. The dashed line represents the true value of *IAR*.

### Posterior prediction of serological data

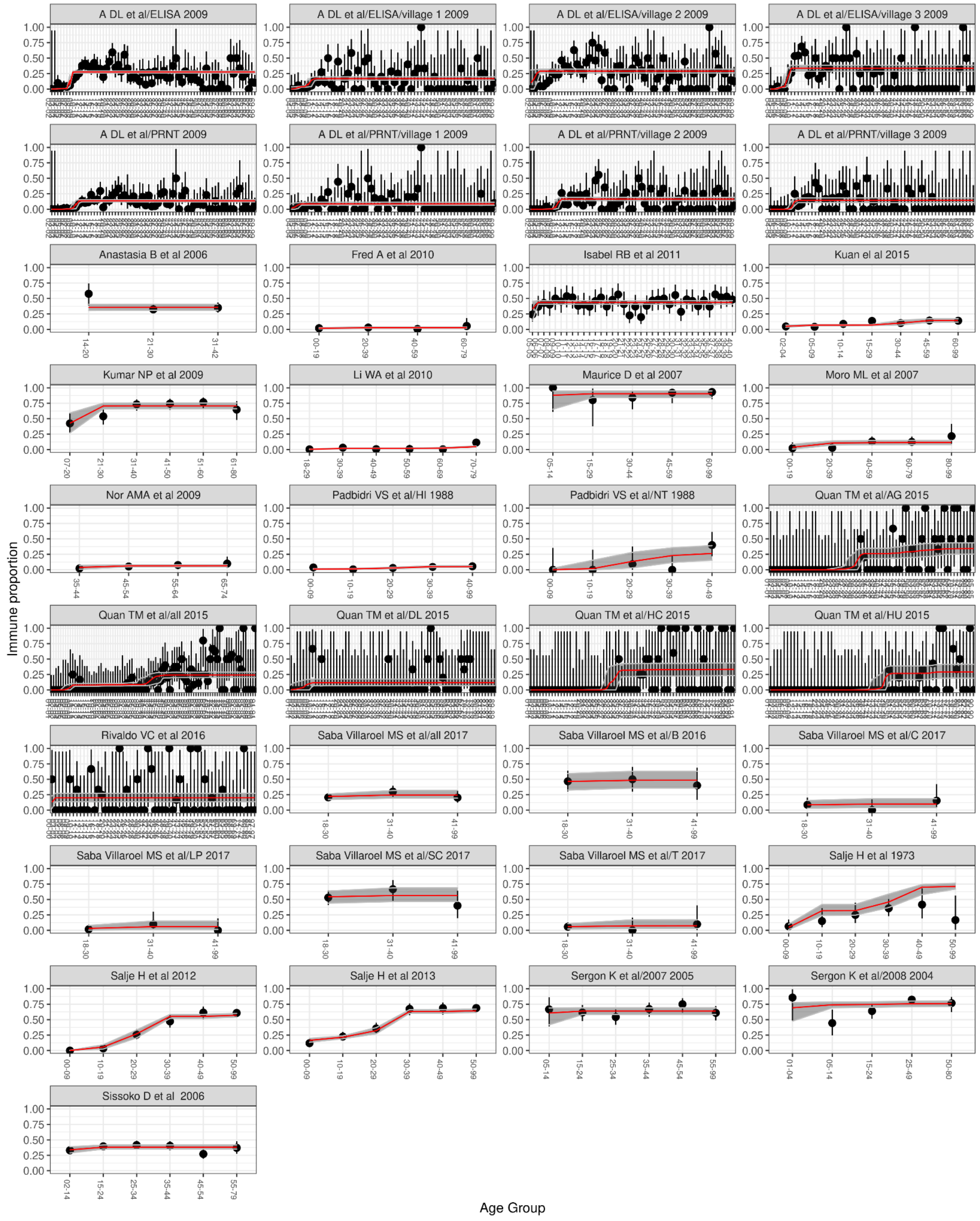


**Figure S10. Datafit from all collated serological data.** The points with their ranges show the true values and 95% CI of the positive proportion for every age group in the simulation data. The red line and the gray area are the median and 95% PPI of ensemble positive proportion for every age group estimated from the 5 models.

### Positive portion for every age group in 2022 in every sites

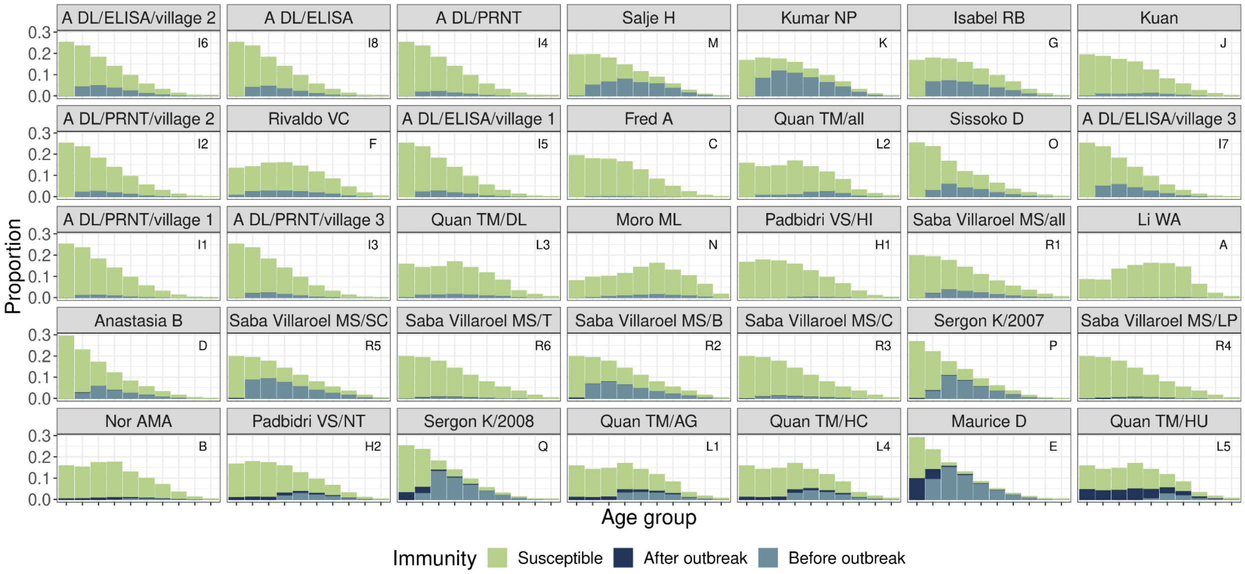


**Figure S11. Positive proportion for every age group of every site before and after the hypothetical outbreak in 2022.** The bar plots for every site show the proportion of immunity in every age group of 10 in the population before (light blue) and after (dark blue) the hypothetical outbreak in 2022. The order of the sites were ranked from the smallest to the largest overall *IAR* in the population (from left to right and from up to down).
