## Supplement Text for "Expected endpoints from future chikungunya vaccine trial sites informed by serological data and modeling"

### Supplemental text

#### Deriving age-stratified susceptible population $S_{a,Y}(\infty)$ after an outbreak

First, we leveraged the relationship between the basic reproduction number ($R_0$), the proportion susceptible in a population before an outbreak ($S(0)$), and the infection attack rate ($IAR$) over the course of an outbreak from a susceptible-infected-recovered (SIR) model [1,2]. This model assumes homogeneous mixing in a closed population and that immunity is lifelong. It predicts that the susceptible population after the outbreak ($S(\infty)$) is

$$S(\infty)=S(0)e^{-R_0(S(0)-S(\infty))}. \quad (1)$$

Then, the infection attack rate is $IAR = S(0) - S(\infty)$. We assumed that the initial prevalence of infection is small, such that $I(0) \approx 0$ and $S(0) \approx 1 - R(0)$, where $R(0)$ is the proportion of the population that has been previously infected and is now immune.

Getting an initial susceptible proportion for the first outbreak can be tricky since there can be outbreaks that happened before the recorded age range. To solve this problem, we used parameter $S_0(0)$ as an average susceptible proportion over all ages of the population at the time before the oldest individuals in the data were born, then this proportion was projected to derive the susceptible population at time $y$ and age $a$ $S_{a,y}(0)$ in the population of the first outbreak.

Since formula (1) only helps us to define the susceptible proportion in a whole population after an outbreak, an extra step was needed to reconstruct the shape of the age-stratified seroprevalence curve. We defined all possible events that can happen to a person in a specific


2

age *a* before and after the outbreak. Let *A* be that a person is at age *a*, *B* is the event that the person is susceptible before an outbreak, and *C* is if the person is still susceptible after the outbreak. *I* is the event that the person is infected during the outbreak. The probability that a person is infected at a certain age *a* and is susceptible before the outbreak is:

$$Pr(I, A, B) = S_a(0) - S_a(\infty) \quad (2),$$

Where $S_a(0)$ and $S_a(\infty)$ are the susceptible proportion at age *a* before and after the outbreak respectively. On the other hand, we could derive this probability differently:

$$Pr(I, A, B) = Pr(I|A, B)\, Pr(A, B) \, (3).$$

We assumed that the transmission of CHIKV is the same for every age group. Therefore, the probability of a person being infected, given that he or she is at age *a* and susceptible before the outbreak is the same for every age, or:

$$Pr(I|A, B) = Pr(I|B) = \frac{Pr(I)}{Pr(B)} \quad (4).$$

From (2), (3), (4) and observing that $Pr(B) = S(0)$, $Pr(C) = S(\infty)$, $Pr(A, B) = S_a(0)$, and $Pr(I) = S(0) - S(\infty)$, we derived this equation:

$$S_a(0) - S_a(\infty) = \left[\frac{S(0) - S(\infty)}{S(0)}\right] S_a(0) \quad (5),$$

which is equivalent to:

$$S_a(\infty) = \frac{S_a(0)\, S(\infty)}{S(0)} \quad (6).$$


3

Therefore, from (1) and (6), by given values of susceptible proportion before the outbreak at a specific age $S_a(0)$ and at the population level $S(0)$, and $R_0$, we could derive the age-specific susceptible proportion after that outbreak $S_a(\infty)$.

Assuming that the outbreak only lasts within a specific year $y$, we derived the age specific susceptible proportions after the outbreak $S_{a,y}(\infty)$ as just stated above. $S_{a,y}(\infty)$ was then projected to the next outbreak time, given that all newborns are susceptible to the infection, to derive $S_{a,y'}(0)$ for the subsequence outbreak at year $y'$. This was then incorporated with the age demographic at that time $pop_{a,y'}$ to derive $S_{y'}(0)$:

$$S_{y'}(0)=\sum_{a=y'-y}^{100} S_{a,y}(\infty)\, pop_{a,y'} \quad (7)$$

We repeated the same process again with the new initial susceptible proportions $S_{a,y'}(0)$ and $S_{y'}(0)$ for the next outbreak. Eventually, after a given number of outbreaks, we could get the age-stratified susceptible proportion $S_{a,y'}(0)$ at study year $Y$. This quantity was used to derive the model's prediction of the proportion positive $P_i$ as stated in the main text.

### Details on curated serosurvey

We applied our approach on published serosurvey data curated from a literature review [3]. The regions of curated study sites consist of South Americas, Asia, and Africa, which are all at high risk of CHIKV emergence. Most of the studies conducted recently, hence they are able to capture the re-emergence era of the virus (2005 - *present*). The studies' quality is varied, ranging from hospital-based with limited recruitment criteria to population-based with systematic sampling in a well-defined population. Details on age groups also varied. Some studies recorded the age of any individuals, others reported results grouped by certain age groups. IgG ELISA is the most common diagnostic test used, though some studies also reported results from HI, IgG IIFT, PRNT or neutralizing tests. For studies that use different tests (H, I) , we ran the model for the results stratified by each test used. The study in Kenya (H) conducted 2 diagnostic tests: IgG ELISA only and PRNT to confirm the ELISA results if there is cross-reactivity with O'nyong-nyong virus. However, only a proportion of IgG ELISA results were confirmed by PRNT. Therefore, we defined the seroprevalence for this dataset by 1) IgG ELISA test results regardless of PRNT results or 2) IgG ELISA test results confirmed by PRNT, and analyzed them individually. Some studies collected samples from multiple sites and we analyzed each site independently, but also ran an extra analysis with seroprevalence of all sites aggregated (L, I, R). For a study in Philippine (M) which had sero surveys for multiple years, we ran the model for all datasets recorded at different years simultaneously (summation of log-likelihood of the all data was used in the Bayesian framework). Details information for each study selected is in the supplementary Table S1. Curated age-stratified seroprevalence data can be found in github [4].

**Table S1. Details information from the selected studies.**

| Index | Author | Country | Adm1 | Time conducted | Time of reported outbreak | Age Range | Samples source | Diagnostic Test |
|---|---|---|---|---|---|---|---|---|
| A | Wei Ang et al. (2017) [5] | Singapore | NA | 2010 | 2008 | 18-79 | residual sera from National Health Survey- a population-based cross-sectional survey among Singapore adult residents. Selection processes are disproportionate stratified sampling and systematic sampling. | ELISA |
| B | Nor Azila Muhammad Azami (2013) [6] | Malaysia | Kuala Lumpur, Selangor, Pahang, and Negeri Sembilan | 2009 | | 35-74 | Participants were from The Malaysian Cohort (TMC) project which is a national project initiated in 2006 to recruit at least 100,000 Malaysians aged 35 and above,where these participants represent various ethnic groups, geographical locations and lifestyles. | IgG ELISA |
| C | Fred Andayi et al (2014) [7] | Djibouti | NA | 11/11/2010 to 02/15/2011 | | every age | 1,045 individuals from 324 households were enrolled randomly. Sources of selected house holds are from the 2009 Hajj Pilgrim database and from the community of health workers (CHW) cognisance list of vulnerable households. | IgG ELISA |

| Index | Author | Country | Adm1 | Time conducted | Time of reported outbreak | Age Range | Samples source | Diagnostic Test |
|---|---|---|---|---|---|---|---|---|
| D | Anastasia Bacci (2015) [8] | Benin | Littoral | 07/2006 to 01/2007 | 2004 | 14-42 | Serum samples are from previous studies on malaria in pregnant women, whether they have fever or not. 352 pregnant women were enrolled at delivery at the Hospital Mother and Child Lanure + Houenoussou Health Center. | IgM and IgG ELISA. samples that were weakly positive for IgG were further tested by indirect immuno-flourescence assay (IIFA) to detect IgG. All positive samples were then confirmed by microneutralization assay (MNTA) |
| E | Maurice Demanou (2010) [9] | Cameroon | Ngehndzen, Ndzeru and Tasaï villages (Western) | 11/2007 | 2006 | over 5 years old | Volunteers | IgM and IgG ELISA |
| F | Rivaldo V. Cunha (2017) [10] | Brazil | Chapada district | 04/2016 | 2014 | every age | Household cluster sampling | IgM and IgG ELISA |
| G | Rodríguez-Barraquer et al. (2015) [11] | India | Tamil Nadu (specifically Chennai) | 06/2011 to 07/2011 | 2006 | 5-40 | Household-based serosurvey | IgG ELISA |
| H | Padbidri et al. (2002) [12] | India | Adaman and Nicobar Islands | 12/1988 to 1/1989 | | every age | 2,401 random samples of local residents across six sites | hemagglutination inhibition/experimental inoculation |
| I | A. Desiree LaBeaud et al. (2015) [13] | Kenya | Kilifi, Kwale, Mombasa | 2009 | 2004 | every age | Samples from a bank serum from another study on polyparasitism in coastal Kenya. | IgG ELISA, PRNT |
| J | Guillermina Kuan et al. (2016) [14] | Nicaragua | Managua | 03-04/2015 and 05/2015 | 2014 | 0-15 and >15 | People in the catchment area of the Health Center Socrates Flores Vivas in District II of Managua. Population estimates in the catchment area were included. | Inhibition ELISA |
| K | Kumar et al. (2011) [15] | India | Kerala (Pathanamthitta, Idukki and Kottaya | 03/2009 - 10/2009 | 2007 | 14-70 | Sample of 360 individuals living in rubber plantation communities | IgG IIFT |


6

| Index | Author | Country | Adm1 | Time conducted | Time of reported outbreak | Age Range | Samples source | Diagnostic Test |
|---|---|---|---|---|---|---|---|---|
| | | | m districts) | | | | | |
| L | Quan et al. (2017) [16] | Vietnam | An Giang, Ho Chi Minh, Dak Lak, Quang Ngai | 2015 | | Every age | residual blood samples from the hospital's outpatients. the patients came to the hospitals without any specific diseases. | IgG ELISA |
| M | Henrik Salje et al. (2015) [17] | Philippines | Cebu | 1973 and 2012 | 1968, 1986, 2012 | Every age | 1973 study: samples are from a cross-sectional study conducted among the general population during parasitological surveys; 2012 study: prospective fever cohort study, randomly sampled individuals ≥6 months of age were enrolled | 1973 study: neutralization assays 2012 study: PCR, PRNT |
| N | Moro et al. (2010) [18] | Italy | Emilia-Romagne | After 2007 | 2007 | Every age | Random sample of 325 villagers from a census list | IgG IFA |
| O | Sissoko et al. (2008) [19] | Mayotte | NA | 11/2006 - 12/2006 | 2005 | > 2 years old | Household-based cross sectional survey | IgM and IgG ELISA |
| P | Sergon et al. (2007) [20] | Comoros | NA | 03/2005 | 2005 | > 5 years old | Cross-sectional survey with systematic sampling | IgM and IgG ELISA |
| Q | Sergon et al. (2008) [21] | Kenya | Lamu Island | 10/2004 | 2004 | > 1 years old | Cross-sectional survey with systematic sampling | IgM and IgG ELISA |
| R | Saba Villaroel et al. (2018) [22] | Bolivia | Santa Cruz, La Paz, Cochabamba, Tarija, and Beni | 12/16 (Beni) and 03/17-04/17 (all others) | 2015 | > 18years old | Blood bank donors | IgG ELISA |

[8] Sambri V, Chippaux J-P, Perrin RX, Massougbodji A, Fievet N, Marchi S, et al. High

Seroprevalence of Chikungunya Virus Antibodies Among Pregnant Women Living in an Urban Area in Benin, West Africa. Am J Trop Med Hyg 2015;92:1133–6. https://doi.org/10.4269/ajtmh.14-0092.



9


10
